# Blood RNA biomarkers for tuberculosis screening in people living with HIV prior to anti-retroviral therapy initiation: A diagnostic accuracy study

**DOI:** 10.1101/2023.06.01.23290783

**Authors:** Tiffeney Mann, Rishi K Gupta, Byron WP Reeve, Gcobisa Ndlangalavu, Aneesh Chandran, Amirtha P Krishna, Claire J Calderwood, Happy Tshivhula, Zaida Palmer, Selisha Naidoo, Desiree L Mbu, Grant Theron, Mahdad Noursadeghi

**Affiliations:** Division of Infection and Immunity, University College London, London, UK; Institute of Health Informatics, University College London, London, UK; DSI-NRF Centre of Excellence for Biomedical Tuberculosis Research; South African Medical Research Council Centre for Tuberculosis Research; Division of Molecular Biology and Human Genetics, Faculty of Medicine and Health Sciences, Stellenbosch University, Cape Town; Department of Clinical Research, Faculty of Infectious and Tropical Diseases, London School of Hygiene & Tropical Medicine, London, UK

**Author notes:** Correspondence Prof Mahdad Noursadeghi, Division of Infection & Immunity, Cruciform Building, University College London, London WC1E 6BT, United Kingdom., Dr Grant Theron, DSI-NRF Centre of Excellence for Biomedical Tuberculosis Research; South African Medical Research Council Centre for Tuberculosis Research; and Division of Molecular Biology and Human Genetics, Faculty of Medicine and Health Sciences, Stellenbosch University, Cape Town, South Africa. Co-first authors (Tiffeney Mann, Rishi K Gupta, Byron WP Reeve). Co-senior authors (Grant Theron, Mahdad Noursadeghi).

## Abstract

**Background:** Undiagnosed tuberculosis (TB) remains a major threat for people living with HIV (PLHIV). Multiple blood transcriptomic biomarkers have shown promise for TB diagnosis. We sought to evaluate their diagnostic accuracy and clinical utility for systematic pre-antiretroviral therapy (ART) TB screening.

**Methods:** We enrolled consecutive adults referred to start ART at a community health centre in Cape Town, South Africa, irrespective of symptoms. Sputa were obtained (using induction if required) for two liquid cultures. Whole-blood RNA samples underwent transcriptional profiling using a custom Nanostring gene-panel. We measured the diagnostic accuracy of seven candidate RNA biomarkers for the reference standard of *Mycobacterium tuberculosis* culture status, using area under the receiver-operating characteristic curve (AUROC) analysis, and sensitivity/specificity at pre-specified thresholds (two standard scores above the mean of healthy controls; Z2). Clinical utility was assessed using decision curve analysis. We compared performance to CRP (threshold ≥5mg/L), World Health Organisation (WHO) four-symptom screen (W4SS) and the WHO target product profile for TB triage tests.

**Results:** A total of 707 PLHIV were included, with median CD4 count 306 cells/mm3. Of 676 with available sputum culture results, 89 (13%) had culture-confirmed TB. The seven RNA biomarkers were moderately to highly correlated (Spearman rank coefficients 0.42-0.93) and discriminated TB culture-positivity with similar AUROCs (0.73-0.80), but none statistically better than CRP (AUROC 0.78; 95% CI 0.72-0.83). Diagnostic accuracy was similar across CD4 count strata, but lower among W4SS-negative (AUROCs 0.56-0.65) compared to W4SS-positive participants (AUROCs 0.75-0.84). The RNA biomarker with highest AUROC point estimate was a 4-gene signature (Suliman4; AUROC 0.80; 95% CI 0.75-0.86), with sensitivity 0.83 (0.74-0.90) and specificity 0.59 (0.55-0.63) at Z2 threshold. In decision curve analysis, Suliman4 and CRP had similar clinical utility to guide confirmatory TB testing, but both had higher net benefit than W4SS. In exploratory analyses, an approach combining CRP (≥5mg/L) and Suliman4 (≥Z2) had sensitivity of 0.80 (0.70-0.87), specificity of 0.70 (0.66-0.74) and higher net benefit than either biomarker alone.

**Interpretation:** RNA biomarkers showed better clinical utility to guide confirmatory TB testing for PLHIV prior to ART initiation than symptom-based screening, but their performance did not exceed that of CRP, and fell short of WHO recommended targets. Interferon-independent approaches may be required to improve accuracy of host-response biomarkers to support TB screening pre-ART initiation.

**Funding:** South African MRC, EDCTP2, NIH/NIAID, Wellcome Trust, NIHR, Royal College of Physicians London.

**Research in Context:** *Evidence before this study:* The World Health Organisation (WHO) commissioned a recent systematic review and individual participant data meta-analysis of tuberculosis (TB) screening strategies among ambulatory people living with HIV (PLHIV). TB is a major cause of morbidity and mortality among PLHIV, particularly among those with untreated HIV and consequent immunosuppression. Importantly, initiation of antiretroviral treatment (ART) for HIV is also associated with increased short-term risk of incident TB, attributed to immune reconstitution inflammatory syndrome, which may in turn potentiate the immunopathogenesis of TB. As a result, in high TB prevalence settings, systematic screening for TB is widely advocated for PLHIV before starting ART. In this context, universal sputum microbiological screening is not economically sustainable, and limited by practical feasibility among those who are not expectorating sputum. Patient stratification to identify those at greater risk of TB is required to target resources for microbiological testing more precisely. For this purpose, the WHO four symptom screen (W4SS) achieved an estimated 84% sensitivity and 37% specificity for pre-ART TB screening. Blood CRP ≥5mg/L offered better performance, estimated at 89% sensitivity and 54% specificity respectively, but still fell short of the WHO target product profile, aiming for ≥90% sensitivity and ≥70% specificity. Blood RNA biomarkers of TB, reflecting interferon (IFN) and tumour necrosis factor-mediated immune responses, have been gaining momentum as potential triage tests for symptomatic and pre-symptomatic TB, but their performance has not been comprehensively evaluated among PLHIV initiating ART. Untreated HIV also drives chronic IFN activity that may compromise the specificity of IFN-dependent biomarkers in this population.

*Added value of this study:* To our knowledge, this is the largest study to date to benchmark the performance of candidate blood RNA biomarkers for unselected and systematic pre-ART TB screening among PLHIV, against contemporary standards and aspirational performance targets. The blood RNA biomarkers showed better diagnostic accuracy and clinical utility to guide confirmatory TB testing for PLHIV than symptom-based screening with W4SS, but their performance did not exceed that of CRP, and they did not achieve WHO recommended targets. The results were comparable for microbiologically confirmed TB at enrolment to the study and for all cases starting TB treatment within six months of enrolment. Blood RNA biomarkers correlated with features of disease severity that might be attributed to either TB or HIV. Accordingly, their discrimination of TB among PLHIV was particularly limited by poor specificity. Diagnostic accuracy was significantly better among people who were symptomatic compared to those who were asymptomatic, further limiting the value of RNA biomarkers in pre-symptomatic TB. Interestingly, blood RNA biomarkers only showed moderate correlation with CRP, suggesting these two measurements provided information on different components of the host response. An exploratory analysis showed that CRP can be combined with the best performing blood RNA signature to provide better clinical utility than achieved by either test alone.

*Implications of all the available evidence:* Our data demonstrate that blood RNA biomarkers do not perform any better than CRP as triage tests for TB among PLHIV prior to ART initiation. Since CRP is already widely available on a low cost point-of-care platform, our findings support further evaluation of the clinical and health-economic impact of CRP-based triage for pre-ART TB screening. An underlying mechanism that limits the diagnostic accuracy of RNA biomarkers for TB among PLHIV prior to ART may be upregulation of interferon signalling in untreated HIV. Since interferon activity underpins upregulated expression of TB biomarker genes, HIV-induced upregulation of interferon-stimulated genes may reduce the specificity of blood transcriptomic biomarkers for TB in this context. These findings highlight a wider need to identify interferon-independent host-response based biomarkers to support disease specific screening of PLHIV pre-ART initiation.

## Introduction

Tuberculosis (TB) continues to be a leading cause of death among people living with HIV (PLHIV) and often remains undiagnosed^1,2^. Prompt diagnosis and treatment initiation are required to reduce mortality in HIV-associated TB (HIV-TB). However, diagnosis can be challenging, with only 56% of estimated incident HIV-TB cases notified in 2019^3^. The World Health Organization (WHO) therefore recommends systematic screening for TB among PLHIV at every healthcare encounter^3^. Since 2011, systematic screening has been based on the WHO four-symptom screen (W4SS) to trigger confirmatory TB testing using culture or molecular assays, and investigate for TB prior to initiation of preventative treatment^3^. W4SS achieves an estimated 84% sensitivity and 37% specificity for TB among outpatients not receiving anti-retroviral therapy (ART)^4^. More recently, WHO have also endorsed measurement of blood C-reactive protein ≥5 mg/L (CRP) for this application, offering similar sensitivity (89%) but higher specificity (54%) than W4SS in an individual participant data meta-analysis^3,4^. However, this still falls short of their target product profile (TPP) which aims to achieve ≥90% sensitivity and ≥70% specificity^5^, necessitating the development of novel approaches with greater accuracy than W4SS or CRP^34^.

Multiple blood transcriptomic biomarkers for TB have shown promise for prediction of incident TB over a 3-6 month interval^6^ and for diagnosis of prevalent TB among symptomatic people^7–9^. Head-to-head analyses have demonstrated areas under the receiver operating characteristic curves (AUROCs) of 0.87-0.91 for the best performing signatures to identify culture-positive TB among symptomatic people self-presenting to healthcare services, irrespective of HIV status^7^. In the context of general population screening, transcriptomic signature AUROCs for prevalent TB have been estimated as 0.63-0.79 in HIV-uninfected people and 0.65-0.88 among PLHIV, with inferior performance among asymptomatic people^8,10^. Signatures are now being translated to near-patient assays, for example using the GeneXpert platform, with promising initial performance among symptomatic individuals^11^. However, data among PLHIV are scarce with only 10 prevalent TB cases in the only previous general population screening study of PLHIV^12^. Moreover, the vast majority of PLHIV in previous studies of both passive and active-case finding were receiving ART. Data from untreated PLHIV are distinctly lacking.

Pre-antiretroviral therapy (ART) initiation is a key timepoint for TB screening, since this frequently represents a nadir of immunocompromise associated with elevated risk of TB both before and during the initial months of ART^13^. Untreated HIV may also be associated with upregulation of interferon signalling^12,14,15^ that may reduce the specificity of blood transcriptomic biomarkers for TB, as also demonstrated during intercurrent respiratory viral infections^16,17^. In this study, we sought to evaluate the diagnostic accuracy and clinical utility of a range of concise candidate blood transcriptomic signatures for TB in a large cohort of PLHIV, in South Africa, prior to commencement of ART. We benchmarked performance against the WHO TPP, along with CRP and W4SS as alternative screening approaches.

## Methods

### Study cohort

Consecutive adults with HIV infection referred to start ART at Kraaifontein Community Health Centre in Cape Town, South Africa, were prospectively enrolled in a parent study evaluating the diagnostic accuracy of sputum Xpert Mtb/Rif Ultra (hereafter, Ultra), irrespective of symptoms. Exclusion criteria included TB treatment within two months or unknown current TB treatment status. In the current study, we included participants who were recruited between 15 May 2017 and 14 December 2020 and who had blood RNA and CRP data available (Figure 1). This study was approved by the Stellenbosch University Faculty of Health Sciences Research Ethics Committee (N14/10/136) and the Western Cape Department of Health, South Africa (WC_2016RP38_944), and is registered on clinicaltrials.gov (NCT03187964). The study is reported in line with STARD guidance^18^.

**Figure 1.**
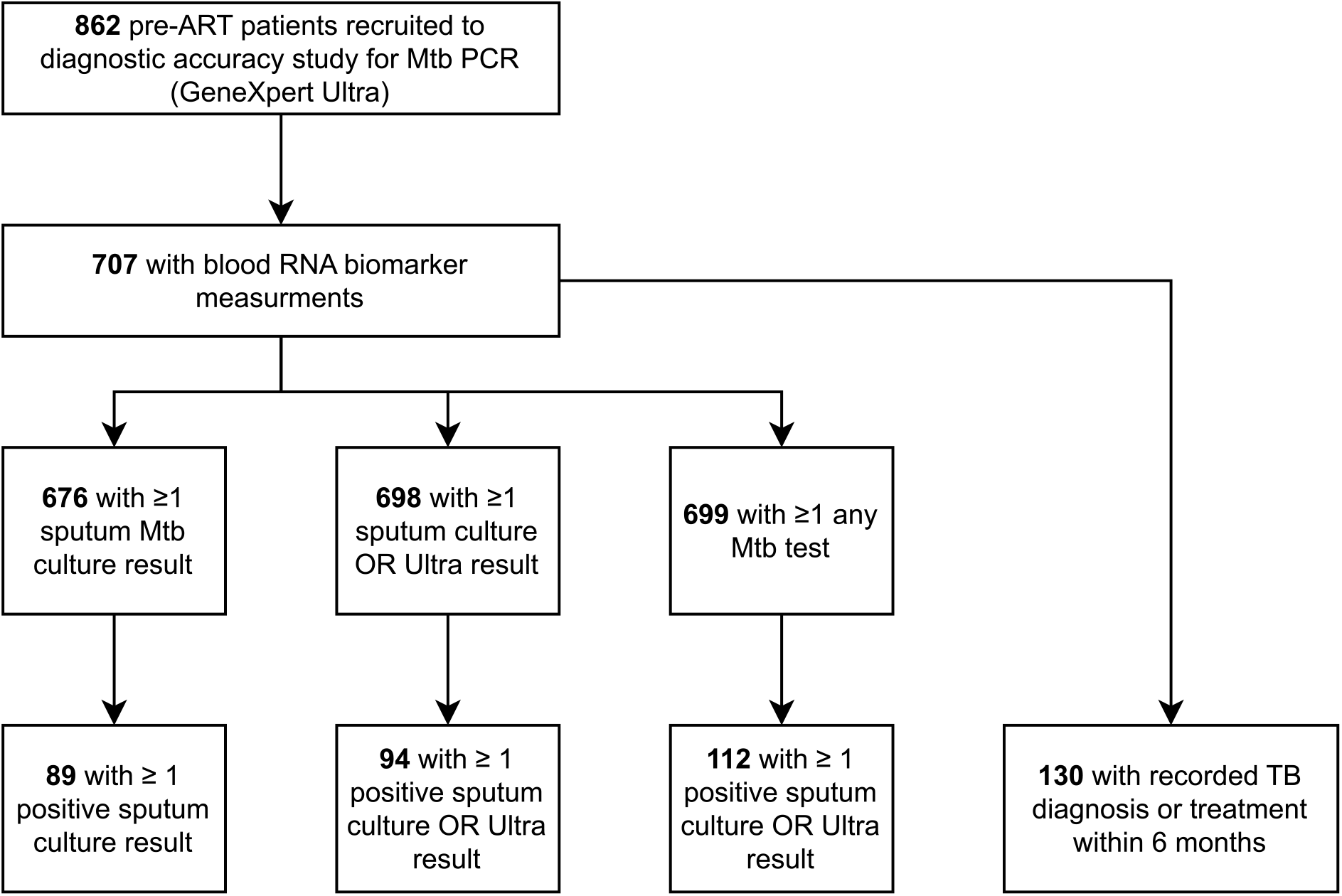
Consort diagram.

Demographic, co-morbidity, symptom and TB treatment data were captured at baseline. CRP was measured in real-time using a point-of-care assay (iChromaII platform, Boditech, South Korea), or a retrospective laboratory assay using stored serum when point-of-care data were unavailable. Three sputum samples were obtained per participant; two underwent smear microscopy and liquid culture and the third sample was tested using Ultra. Expectoration was attempted for at least one sample. If this was not sufficient, sputum induction was performed using nebulised hypertonic saline. Blood RNA was collected in Tempus tubes (Ambion, Life Technologies) and preserved at -80ºC. Urine samples were collected and tested using Ultra and Determine LF-LAM (Abbott, South Africa). TB diagnoses and treatment data after enrolment were obtained through linkage to routinely collected health record data held at the Western Cape Provincial Health Data Centre using a deterministic algorithm^19^.

The reference standard for primary analyses was sputum liquid culture status for *Mycobacterium tuberculosis complex*. Secondary reference standards included: (1) sputum culture *or* Ultra positivity (excluding trace positive results); (2) any positive TB test, including urine LAM or Ultra (excluding sputum Ultra trace positive results); and (3) recorded TB diagnosis or TB treatment in study or linkage data within six months of enrolment. Participants with missing blood RNA or outcome data were excluded for each analysis.

Our sample size calculation was based on achieving a minimum AUROC of 0.8, equivalent to that of CRP^4^ with a 95% lower confidence interval bound of 0.75, requiring 700 participants with a minimum TB prevalence rate of 10% (Supplementary Figure 1)^20^.

### Sample processing and Nanostring

Peripheral blood RNA samples were extracted using the Tempus Spin RNA Isolation kit (Ambion, Life Technologies) and the Turbo DNA-free Kit (Invitrogen). RNA integrity scores were determined using the Agilent Tape Station (Agilent). Transcriptional profiling of 300ng of blood RNA was performed using a custom gene panel on the Nanostring platform (Nanostring Technologies). The gene panel was designed to include 23 TB genes encompassing seven candidate RNA signatures for TB, limited to concise signatures of ≤11 genes that performed well in our previous head-to-head analyses^7,21^. The signatures included were Suliman4, RISK6, Sweeney3, Giddon3, BATF2, Roe3 and Zak11. Signatures are referred to with a prefix of the first-author’s surname from the original publication where the signature was derived, and a suffix of the number of component genes, with the exceptions of BATF2 (a single transcript) and RISK6 (as named by the original investigators)^22^.

Nanostring data were analysed on the nCounter Analyser (Nanostring Technologies) using 555 field of view, as per manufacturer’s instructions. Quality control and gene expression normalisation were performed with nSolver Analysis software (version 4.0.70). Gene expression values were log2-transformed and then normalised by subtracting the log2-expression of a housekeeping gene (*GAPDH*). Blood RNA signature Z-scores were calculated by standardising scores for each signature to the mean and standard deviation of blood samples (N=105) from a healthy control population of individuals with latent TB^23^, also measured using the same Nanostring codeset.

Reference RNA samples (Universal Human Reference RNA, Agilent) were included in each Nanostring run to facilitate quality control. These demonstrated minimal coefficients of variation for each gene, supporting reproducibility of measurements (Supplementary Figure 2). We also examined the discrimination of TB and non-TB cases using Nanostring measurements of the seven candidate RNA signatures in 59 paired RNA samples from our previously reported presumptive TB cohort^7^, to ensure that we could reproduce the results derived from RNAseq data (Supplementary Figure 3).

Principal component analyses revealed systematic differences in reference RNA data by manufacturing codeset batch (Supplementary Figure 4). We therefore performed batch correction by codeset manufacturing batch, using the *ComBat* function from the *sva* package in R^24^. Distributions of target genes differed by batch to a varying degree for each probe prior to batch correction (Supplementary Figure 5). These differences resolved after correction (Supplementary Figure 6-7).

### Data analysis

Analyses were conducted R (version 4.0.2). Blood RNA signature scores were calculated from processed Nanostring data as reported previously (Supplementary Table 1)^7,21^. For the Roe3 signature, we sought to simplify our approach to calculating signature scores to promote generalisability. We examined whether a geometric mean calculation for the three component genes would perform as well as the original support vector machine approach in our previous incipient^21^ and presumptive TB^7^ datasets. Since the simplified (geometric mean) approach performed similarly, this method was used in all subsequent analyses (Supplementary Figure 8).

We quantified discrimination for each biomarker by constructing receiver operating characteristic (ROC) curves and calculating areas under the curves (AUROCs) using the *pROC* package, with 95% confidence intervals using the DeLong method^25^. We also compared AUROCs for each RNA signature to CRP using paired DeLong tests, with adjustment for multiple testing using a Benjamini-Hochberg correction^25^. Sensitivities, specificities and predictive values for each RNA signature were calculated using pre-specified cut-offs of two standard scores above the mean of the healthy control population (Z2).

Subgroup analyses were performed by stratifying by W4SS status (presence of any of fever, cough, weight loss or sweats) and CD4 count (<200 cells/mm^3^ *vs*. ≥200 cells/mm^3^). We also compared discrimination between participants with a recorded TB diagnosis or treatment within six months of enrolment and those who remained TB-free for this period, stratified by sputum culture, to assess if accuracy varied according to sputum culture status. We compared discrimination for each signature between subgroups using unpaired Delong tests, with adjustment for multiple testing as before.

Correlation between RNA signatures, CRP and physiological indices of disease severity were examined using scatterplots and Spearman rank correlation coefficients. We also examined factors associated with higher RNA signature scores using multivariable linear regression, with restricted cubic splines for continuous variables to account for potential non-linear associations.

In order to investigate the clinical utility of candidate TB screening strategies, decision curve analysis was performed using the *rmda* package^26^, as described previously^27^. Briefly, decision curve analysis determines the ‘net benefit’ of diagnostic approaches, in comparison to intervening for all or no participants. Net benefit reflects the true positive rate minus false positive rate weighted by the cost-benefit ratio across a range of threshold probabilities which will trigger a decision, as a surrogate measure of the range of cost-benefit ratios. The “intervention” in the context of a TB screening test is the offer of confirmatory testing (for example using sputum culture), and the threshold probabilities reflect the minimum probability of disease at which further investigation would be triggered. The ideal approach has the highest net-benefit across a clinically relevant threshold probability range. The net-benefit of using the best performing RNA biomarker (at a threshold of Z2) to guide confirmatory testing was assessed, compared to alternative strategies of confirmatory testing for all, confirmatory testing for none, and confirmatory testing guided by CRP (cut-off ≥5mg/L as recommended in WHO guidance^3,4^) or W4SS.

We also performed a range of exploratory analyses. First, we examined whether an optimised HIV-TB RNA signature could be derived by temporally splitting the cohort into development (75%) and validation (25%) sets. We then ranked the 23 measured transcripts by AUROC for TB culture status in the development set and examined whether iteratively adding genes improved discrimination in the held-out validation set. We used a range of approaches to combine individual genes to calculate overall scores, including simple calculations (geometric means or disease risk scores^28^), and multivariable models trained on the development set (logistic regression and support vector machines). Second, we used the same development/validation split to assess whether a multivariable model including the most discriminating RNA signature, CRP and clinical predictors (number of W4SS symptoms, haemoglobin, CD4 count and body mass index) may further improve performance. For this, we used a multivariable logistic regression approach with restricted cubic splines to model potential non-linear associations. Finally, we examined whether an approach combining CRP (≥5mg/L) *and* the most discriminating RNA biomarker (Z-score ≥2) may offer better net benefit to guide confirmatory testing in decision curve analysis.

### Sensitivity analyses

We explored the effect of alternative reference standard definitions, as described above, in sensitivity analyses. We also examined an alternative approach to Nanostring data batch-correction, by normalising probe-level data to the mean of the reference RNA samples for each batch.

## Results

### Overview of study cohort

A total of 862 participants were recruited to the parent study during the study period. Of these, 707 (82%) had blood RNA and CRP data available and were included in the analysis (Figure 1, Table 1). There were no systematic differences between included and excluded participants (Supplementary Table 2). Of the included study population (n=707), median age was 32 years (interquartile range [IQR] 27-39), 407 (56%) were female and median CD4 cell count was 306 cells/mm^3^ (IQR 184-486). A total of 406 (57%) presented with at least one of the symptoms comprising the W4SS, while 633 (90%) of the included cohort had two sputum culture results available. Of 676 participants with at least one sputum culture result, 89 (13.2%) were positive, while 65/699 (9.3%) with available Ultra results were positive. 130/707 (18%) of participants had a recorded TB diagnosis or treatment within 6 months of enrolment; 84% and 89% of these cases were within 4 and 8 weeks, respectively (Supplementary Figure 9). A total of 11/107 (10.2%) of those with known site of disease were extra-pulmonary.

**Table 1.**
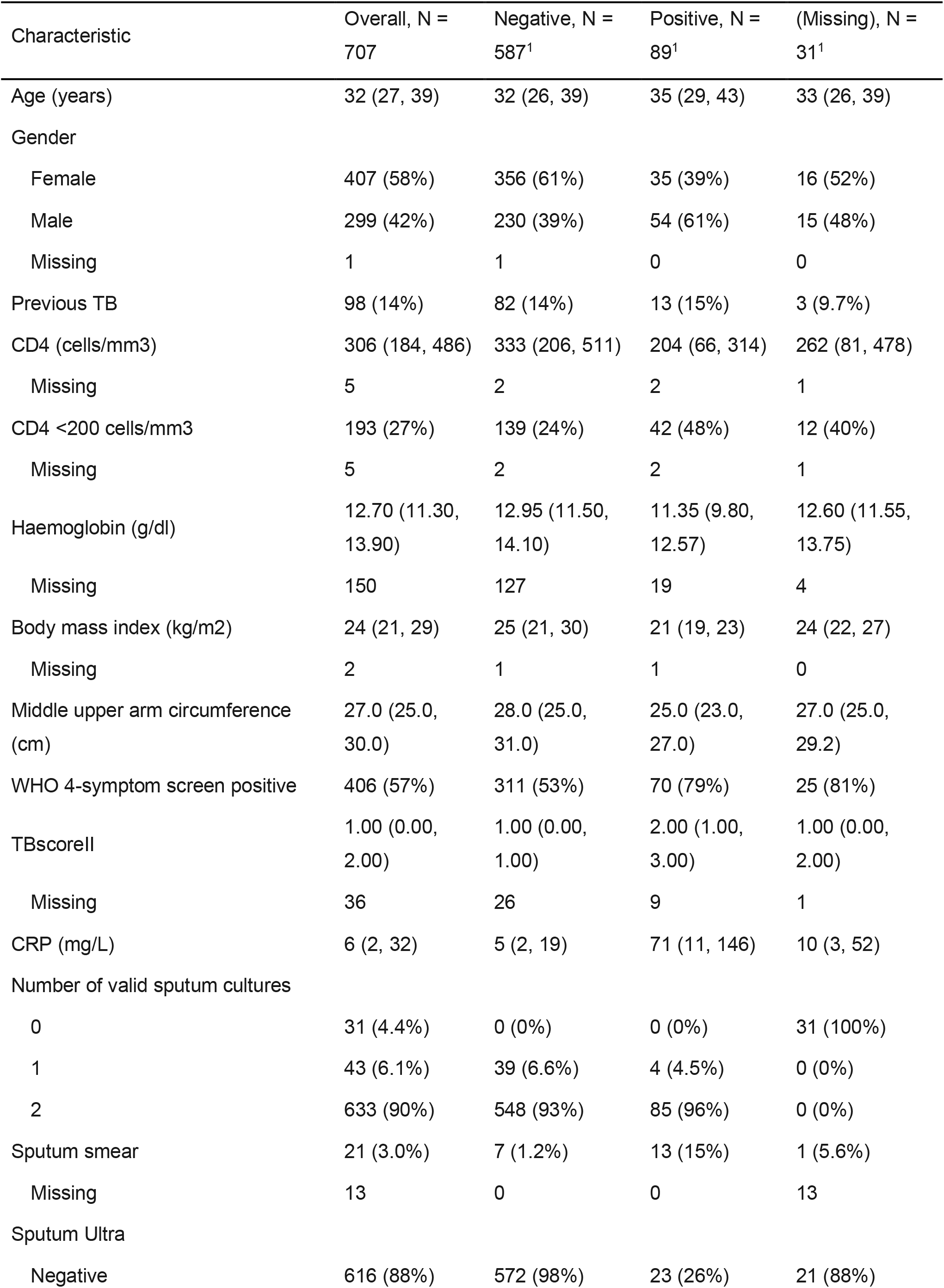
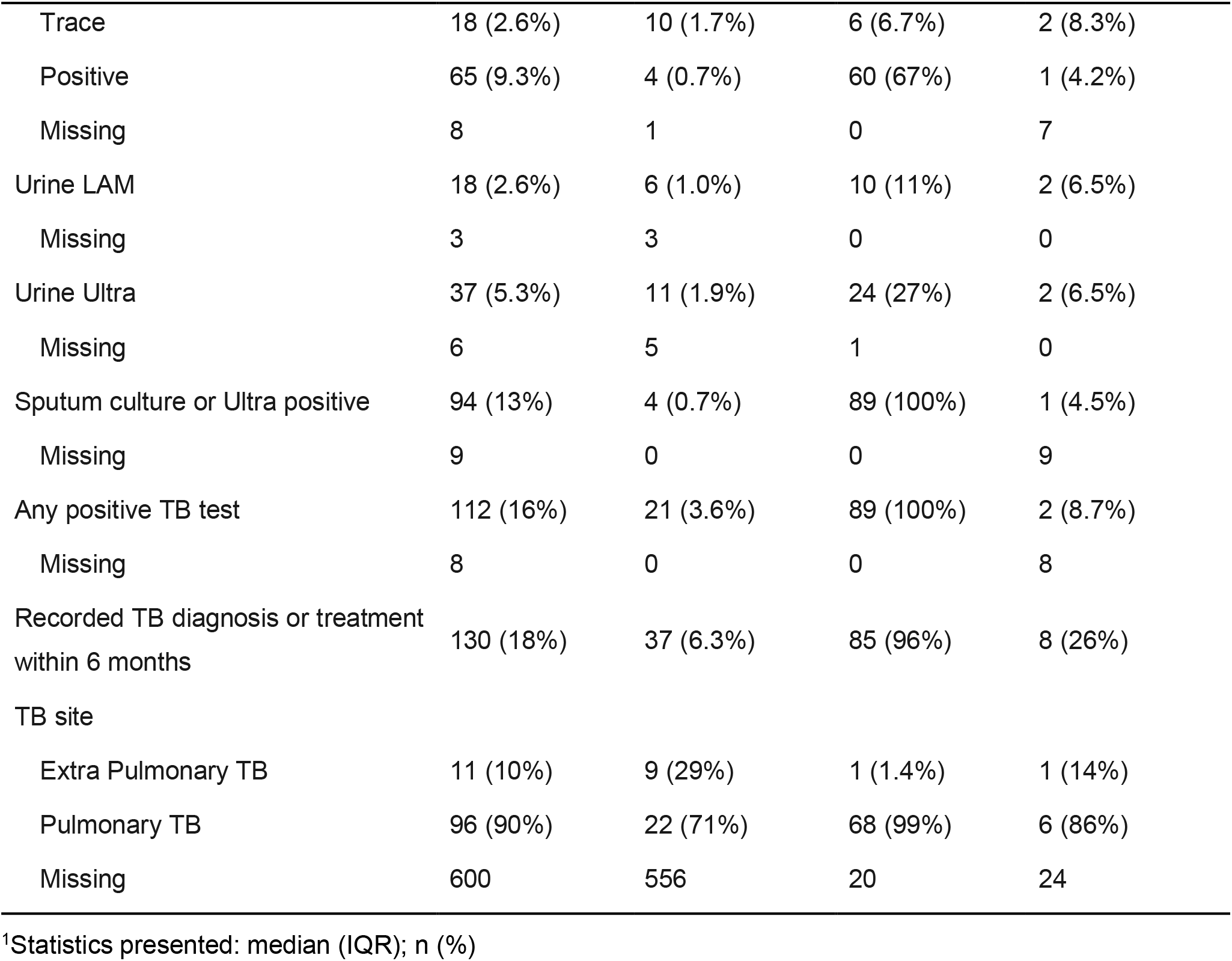
Baseline characteristics of the included study cohort, stratified by culture status.

| Characteristic | Overall, N =<br>707 | Negative, N =<br>587<sup>1</sup> | Positive, N =<br>89<sup>1</sup> | (Missing), N =<br>31<sup>1</sup> |
| --- | --- | --- | --- | --- |
| Age (years) | 32 (27, 39) | 32 (26, 39) | 35 (29, 43) | 33 (26, 39) |
| Gender |  |  |  |  |
| Female | 407 (58%) | 356 (61%) | 35 (39%) | 16 (52%) |
| Male | 299 (42%) | 230 (39%) | 54 (61%) | 15 (48%) |
| Missing | 1 | 1 | 0 | 0 |
| Previous TB | 98 (14%) | 82 (14%) | 13 (15%) | 3 (9.7%) |
| CD4 (cells/mm3) | 306 (184, 486) | 333 (206, 511) | 204 (66, 314) | 262 (81, 478) |
| Missing | 5 | 2 | 2 | 1 |
| CD4 <200 cells/mm3 | 193 (27%) | 139 (24%) | 42 (48%) | 12 (40%) |
| Missing | 5 | 2 | 2 | 1 |
| Haemoglobin (g/dl) | 12.70 (11.30,<br>13.90) | 12.95 (11.50,<br>14.10) | 11.35 (9.80,<br>12.57) | 12.60 (11.55,<br>13.75) |
| Missing | 150 | 127 | 19 | 4 |
| Body mass index (kg/m2) | 24 (21, 29) | 25 (21, 30) | 21 (19, 23) | 24 (22, 27) |
| Missing | 2 | 1 | 1 | 0 |
| Middle upper arm circumference (cm) | 27.0 (25.0,<br>30.0) | 28.0 (25.0,<br>31.0) | 25.0 (23.0,<br>27.0) | 27.0 (25.0,<br>29.2) |
| WHO 4-symptom screen positive | 406 (57%) | 311 (53%) | 70 (79%) | 25 (81%) |
| TBscoreII | 1.00 (0.00,<br>2.00) | 1.00 (0.00,<br>1.00) | 2.00 (1.00,<br>3.00) | 1.00 (0.00,<br>2.00) |
| Missing | 36 | 26 | 9 | 1 |
| CRP (mg/L) | 6 (2, 32) | 5 (2, 19) | 71 (11, 146) | 10 (3, 52) |
| Number of valid sputum cultures |  |  |  |  |
| 0 | 31 (4.4%) | 0 (0%) | 0 (0%) | 31 (100%) |
| 1 | 43 (6.1%) | 39 (6.6%) | 4 (4.5%) | 0 (0%) |
| 2 | 633 (90%) | 548 (93%) | 85 (96%) | 0 (0%) |
| Sputum smear | 21 (3.0%) | 7 (1.2%) | 13 (15%) | 1 (5.6%) |
| Missing | 13 | 0 | 0 | 13 |
| Sputum Ultra |  |  |  |  |
| Negative | 616 (88%) | 572 (98%) | 23 (26%) | 21 (88%) |
| Trace | 18 (2.6%) | 10 (1.7%) | 6 (6.7%) | 2 (8.3%) |
| Positive | 65 (9.3%) | 4 (0.7%) | 60 (67%) | 1 (4.2%) |
| Missing | 8 | 1 | 0 | 7 |
| Urine LAM | 18 (2.6%) | 6 (1.0%) | 10 (11%) | 2 (6.5%) |
| Missing | 3 | 3 | 0 | 0 |
| Urine Ultra | 37 (5.3%) | 11 (1.9%) | 24 (27%) | 2 (6.5%) |
| Missing | 6 | 5 | 1 | 0 |
| Sputum culture or Ultra positive | 94 (13%) | 4 (0.7%) | 89 (100%) | 1 (4.5%) |
| Missing | 9 | 0 | 0 | 9 |
| Any positive TB test | 112 (16%) | 21 (3.6%) | 89 (100%) | 2 (8.7%) |
| Missing | 8 | 0 | 0 | 8 |
| Recorded TB diagnosis or treatment<br>within 6 months | 130 (18%) | 37 (6.3%) | 85 (96%) | 8 (26%) |
| TB site |  |  |  |  |
| Extra Pulmonary TB | 11 (10%) | 9 (29%) | 1 (1.4%) | 1 (14%) |
| Pulmonary TB | 96 (90%) | 22 (71%) | 68 (99%) | 6 (86%) |
| Missing | 600 | 556 | 20 | 24 |

<sup>1</sup>Statistics presented: median (IQR); n (%)

### Diagnostic accuracy of RNA signatures for culture-positive TB

The seven RNA biomarkers had similar discrimination for sputum culture status, with AUROCs ranging from 0.73 for Zak11 (95% CI 0.68-0.79) to 0.80 for Suliman4 (95% CI 0.75-0.86) (Figure 2). All of the RNA biomarkers had statistically equivalent discrimination to CRP in pairwise tests (CRP AUROC 0.78; 95% CI 0.72-0.83; Table 2). Using Z2 cut-offs, none of the signatures met the WHO-recommended minimum sensitivity of 90% and specificity of 70% for a triage test (Table 2). Suliman4, the RNA signature with the highest AUROC point estimate, had sensitivity and specificity of 0.83 (0.74-0.9) and 0.59 (0.55-0.63), respectively. Most signatures had higher sensitivity than specificity at Z2 cut-offs, with 28-74% of participants having a score above the Z2 threshold. Positive predictive values ranged from 16-27%, with negative predictive values 92-97%. By comparison, CRP had sensitivity of 0.85 (0.77-0.91) and specificity 0.48 (0.44-0.52) at the primary cut-off of ≥5mg/L, with 56% of participants having a positive result. For Suliman4 at Z2 threshold, the numbers needed to test with confirmatory testing were 4.3 (3.5-5.3) and 23.9 (14.8-39.2) among Suliman4-positive and negative participants. For CRP, numbers needed to test were similar at 5 (4.1-6.2) and 22.7 (13.5-38.6) among CRP-positive and negative individuals, respectively.

**Figure 2.**
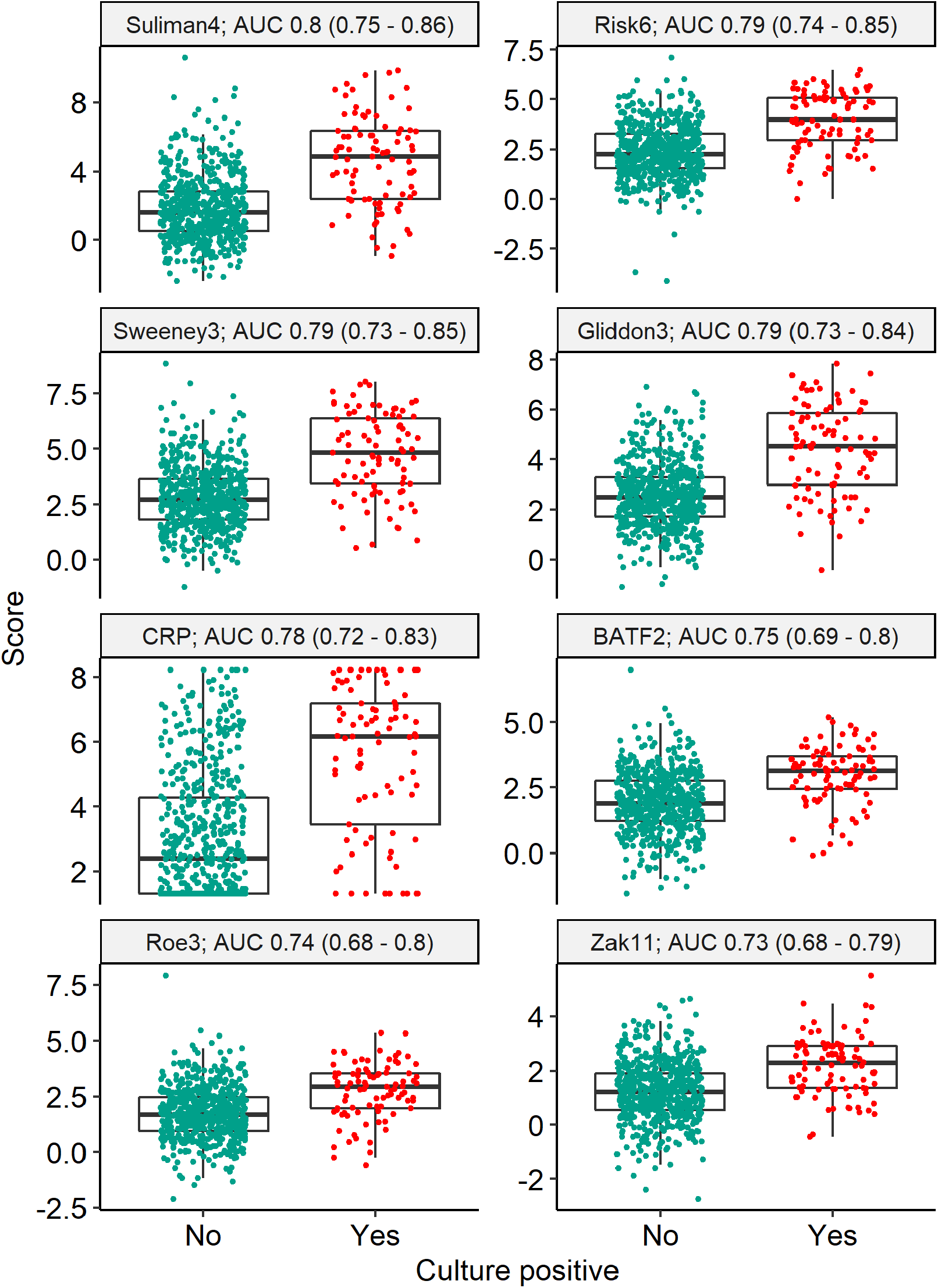
Blood RNA biomarker discrimination of Mtb sputum culture positivity. Scores and discrimination of RNA signatures and CRP for primary outcome of sputum culture positivity (n = 676 participants). Scores are shown as Z-scores for RNA signatures, and log-2 transformed CRP (mg/L). Discrimination presented as area under the receiver operating characteristic curve (AUC), with 95% confidence intervals.

**Table 2.**
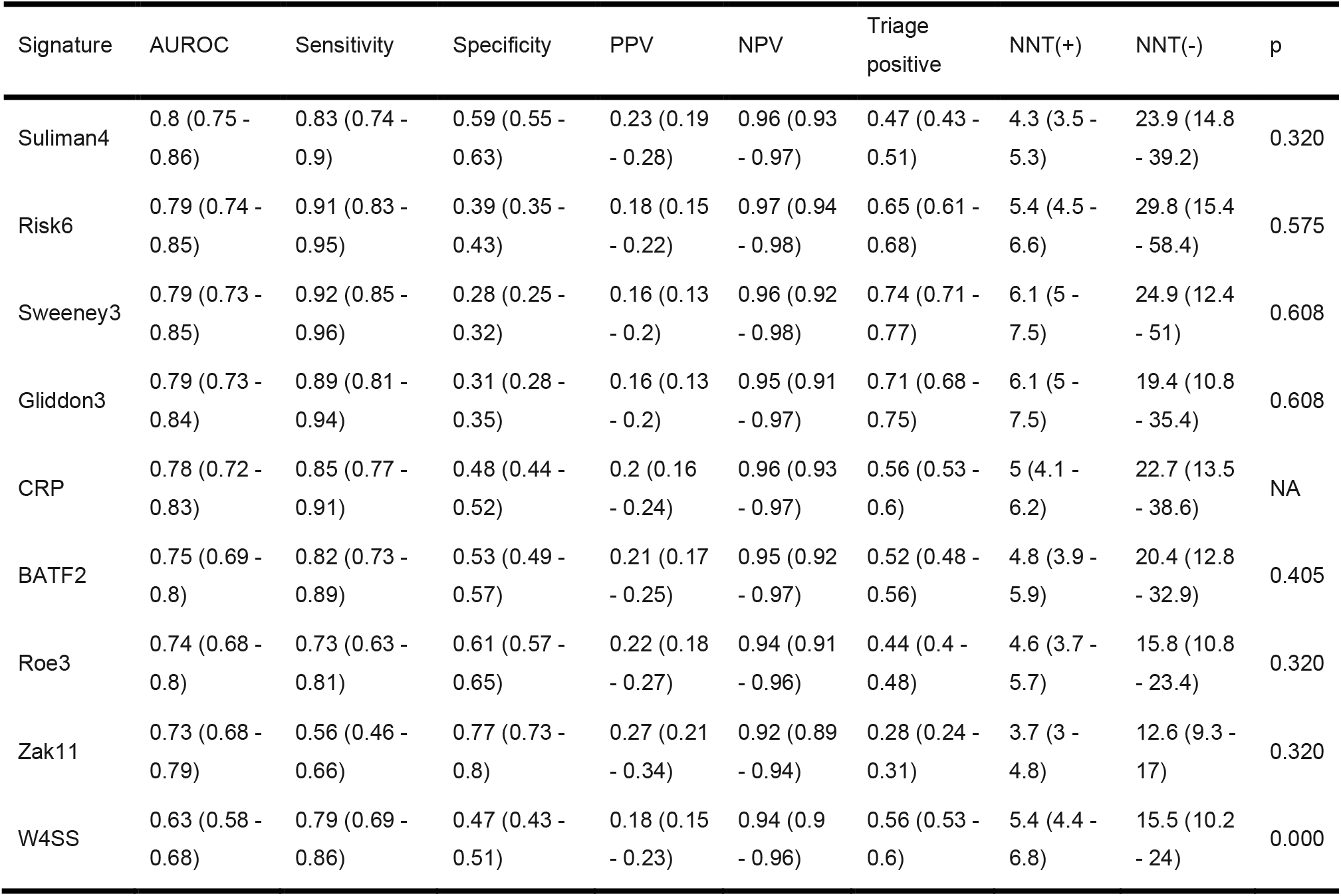
Blood RNA biomarker and CRP performance metrics for discrimination of sputum culture status. P values indicate pairwise comparisons to CRP, with multiple testing correction (n = 676 participants).

In subgroup analyses, RNA signatures and CRP were less discriminating among W4SS-negative (AUROCs 0.56-0.65), compared to W4SS-positive participants (AUROCs 0.75-0.84; Table 3). There were no differences in discrimination when stratified by CD4 count or sputum culture status of TB cases.

**Table 3.** Blood RNA biomarker and CRP performance metrics for discrimination of TB status, stratified by selected subgroups. Accuracy shown for primary outcome of sputum culture status as area under the receiver operating characteristic curve with 95% confidence intervals. Reference standard for W4SS and CD4 strata is sputum culture, as per primary reference standard. Reference standard for the sputum culture strata is based on TB diagnosis or treatment recorded within six months of enrolment. P values indicate comparisons between strata for each signature.

| Signature | W4SS-positive | W4SS-negative | p |
| --- | --- | --- | --- |
| BATF2 | 0.78 (0.72 - 0.84) | 0.59 (0.43 - 0.74) | 0.029 |
| CRP | 0.81 (0.76 - 0.87) | 0.56 (0.42 - 0.7) | 0.003 |
| Gliddon3 | 0.83 (0.77 - 0.89) | 0.58 (0.44 - 0.72) | 0.003 |
| Risk6 | 0.83 (0.78 - 0.89) | 0.59 (0.46 - 0.72) | 0.003 |
| Roe3 | 0.77 (0.71 - 0.83) | 0.57 (0.41 - 0.74) | 0.029 |
| Suliman4 | 0.84 (0.79 - 0.89) | 0.6 (0.46 - 0.74) | 0.003 |
| Sweeney3 | 0.84 (0.79 - 0.9) | 0.56 (0.42 - 0.7) | 0.002 |
| Zak11 | 0.75 (0.68 - 0.81) | 0.65 (0.51 - 0.78) | 0.193 |
| Signature | CD4 <200 cells | CD4 ≥200 cells | p |
| BATF2 | 0.76 (0.68 - 0.84) | 0.7 (0.62 - 0.79) | 0.482 |
| CRP | 0.84 (0.77 - 0.91) | 0.7 (0.61 - 0.78) | 0.112 |
| Gliddon3 | 0.83 (0.75 - 0.9) | 0.72 (0.63 - 0.81) | 0.188 |
| Risk6 | 0.83 (0.75 - 0.9) | 0.73 (0.65 - 0.82) | 0.188 |
| Roe3 | 0.75 (0.66 - 0.83) | 0.7 (0.61 - 0.79) | 0.512 |
| Suliman4 | 0.84 (0.76 - 0.92) | 0.75 (0.67 - 0.83) | 0.188 |
| Sweeney3 | 0.84 (0.76 - 0.91) | 0.73 (0.65 - 0.82) | 0.188 |
| Zak11 | 0.71 (0.62 - 0.8) | 0.74 (0.65 - 0.82) | 0.667 |
| Signature | Culture-positive TB | Culture-negative TB | p |
| BATF2 | 0.75 (0.7 - 0.81) | 0.69 (0.61 - 0.77) | 0.447 |
| CRP | 0.79 (0.73 - 0.85) | 0.79 (0.72 - 0.86) | 0.978 |
| Gliddon3 | 0.8 (0.74 - 0.86) | 0.76 (0.68 - 0.84) | 0.549 |
| Risk6 | 0.8 (0.75 - 0.86) | 0.75 (0.67 - 0.83) | 0.447 |
| Roe3 | 0.75 (0.69 - 0.81) | 0.69 (0.61 - 0.77) | 0.447 |
| Suliman4 | 0.82 (0.76 - 0.87) | 0.79 (0.72 - 0.86) | 0.689 |
| Sweeney3 | 0.8 (0.74 - 0.85) | 0.7 (0.61 - 0.78) | 0.361 |
| Zak11 | 0.74 (0.69 - 0.8) | 0.67 (0.6 - 0.75) | 0.447 |

### Associations between RNA scores, indices of HIV/TB severity and CRP

The seven RNA biomarkers were moderately to highly correlated (Spearman rank coefficients 0.42-0.93; Supplementary Figure 10). Correlation was also observed between CRP and RNA biomarkers (Spearman rank coefficients 0.23-0.61), though this tended to be weaker than that observed between the RNA biomarkers.

To examine whether RNA signature scores were associated with HIV/TB severity, we plotted scatterplots for Suliman4 (the signature with the highest AUROC point estimate) with clinical measures of disease severity, stratified by TB status (Supplementary Figure 11). Higher Suliman4 scores were associated with lower BMI, CD4 count, haemoglobin and middle upper arm circumference, along with higher TBScoreII^29^ and respiratory rate among participants with and without TB (Supplementary Table 2). Among those with TB, higher Suliman4 scores were associated with higher smear grade and lower time to positivity in liquid culture. In multivariable linear regression, number of symptoms, BMI, CD4 count, haemoglobin, respiratory rate and sputum culture status were independently associated with higher Suliman4 scores (Supplementary Figure 12).

### Clinical utility

In decision curve analysis, Suliman4 with a Z2 cut-off to guide confirmatory testing had higher net benefit than an approach of confirmatory testing for all when the threshold probability exceeded ∼4% (equivalent to a number willing to test with confirmatory testing of up to ∼24 people per true TB case detected; Figure 3; Supplementary Table 3). Using CRP ≥5mg/L had similar, albeit slightly lower net benefit to Suliman4. There was a larger incremental net benefit for Suliman4 with increasing threshold probabilities (i.e. when the number willing to test is lower). Both Suliman4 and CRP had higher net benefit than W4SS, which itself surpassed a confirmatory testing for all approach above threshold probabilities of ∼6% (equivalent to a number willing to test with confirmatory testing of up to ∼15 people per true TB case detected).

**Figure 3.**
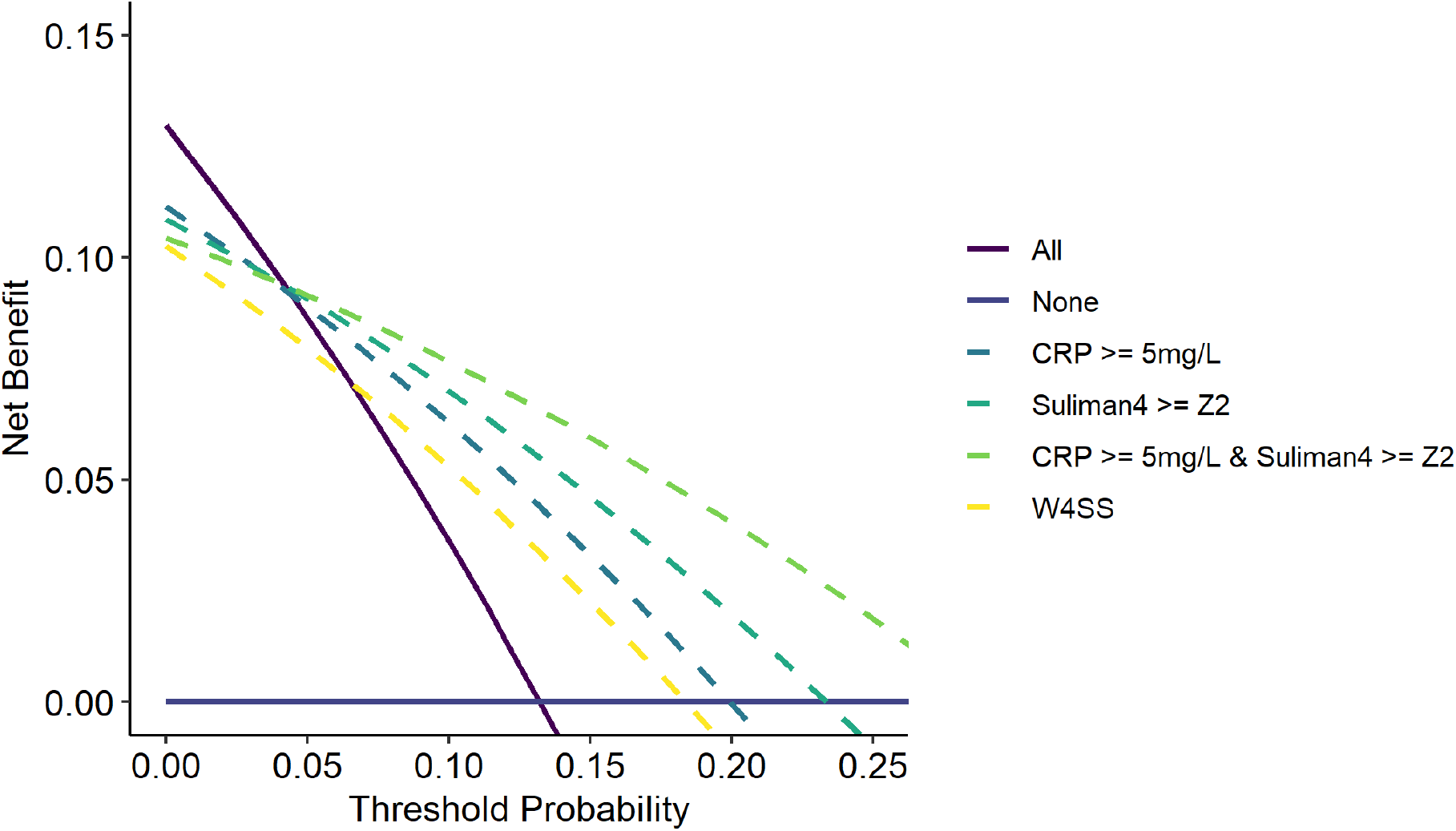
Decision curve analysis for alternative triage strategies to trigger confirmatory investigations for TB. Net benefit (true positive rate - false positive rate weighted by cost:benefit ratio) for investigate all and investigate none approaches across the range of threshold probabilities that a service may use to trigger confirmatory investigations for TB is compared to that of decisions to investigate triggered by each of triage strategies indicated: CRP (≥5 and ≥10mg/L), Suliman4 (Z2), symptoms (W4SS) and using an exploratory approach of CRP>=5 and Suliman4 >=Z2.

### Exploratory analyses

Our forward search to identify an optimised RNA signature for HIV-TB did not lead to significantly improved discrimination for TB culture status in the temporal validation set, when using simple calculations or multivariable models to combine individual gene expression values (Supplementary Figure 13, Supplementary Table 4). A multivariable model trained on the development set incorporating Suliman4, CRP and clinical predictors (number of W4SS symptoms, haemoglobin, CD4 count and body mass index) also did not lead to significant improvement in discrimination, with AUROC 0.81 (0.71 - 0.91) in the validation set. Our exploratory approach of combining CRP (≥5mg/L) and Suliman4 (≥Z2) had sensitivity of 0.80 (0.70 - 0.87) and specificity of 0.70 (0.66 - 0.74). In decision curve analyses, this approach had slightly higher net benefit than Suliman4 alone, with an incrementally greater net benefit at higher threshold probabilities (Figure 5).

### Sensitivity analyses

Our sensitivity analyses using alternative reference standard definitions for TB and an alternative approach to Nanostring data batch-correction did not lead to any substantial difference in the main results (Supplementary Figures 15-17).

## Discussion

To our knowledge, this is the first study to examine the diagnostic accuracy and clinical utility of a range of promising blood RNA signatures for the application of systematic HIV-TB screening prior to ART initiation. We found that the seven candidate signatures had similar diagnostic accuracy for culture-positive TB and were moderately to highly correlated, supporting previous analyses. However, none of the candidate signatures met the WHO target product profile criteria for a triage test and performance did not exceed that of CRP. The RNA signature with the highest point estimate was Suliman4; both Suliman4 and CRP had superior clinical utility to guide confirmatory testing compared to W4SS. Signature accuracy appeared independent of CD4 count and the sputum culture status of TB cases. However, accuracy was lower among W4SS-negative participants for all signatures and CRP, suggesting inferior performance in the absence of symptomatic disease.

While most RNA signatures either met or approached the WHO triage test sensitivity target of 90% at the Z2 cut-off, specificity was generally below the 70% target. This lack of specificity may reflect upregulation of interferon activity that we have previously shown underpins expression of TB biomarker genes^6^. Such upregulation of interferon activity may be driven by untreated HIV itself^12,14,15^, and/or other opportunistic infections. Notably, higher RNA signature scores were associated with indices of HIV severity (including lower BMI, haemoglobin and CD4 count) in univariable and multivariable analyses, further supporting this hypothesis. Collectively, these findings highlight the need to develop interferon-independent host response biomarkers for TB screening among PLHIV.

In our clinical utility analyses, we showed that both Suliman4 and CRP had higher net benefit than confirmatory testing for all if the health service is willing to perform up to approximately 22 confirmatory tests per true TB case diagnosed. If the health service can perform more confirmatory tests than this, then our analysis suggests a confirmatory testing for all approach may be preferable. Of note, our exploratory approach combining CRP (≥5mg/L) and Suliman4 (≥Z2) showed higher net benefit than either biomarker alone, with largely preserved sensitivity of 80% and improved specificity of 70%. This finding reflects that blood RNA biomarkers and CRP had weak to moderate correlation, they may therefore provide orthogonal information. Combining both has the potential to improve specificity, however such an approach would incur additional cost, making it highly unlikely to achieve the WHO target price for a triage test of <$2. Since CRP testing is already widely available, including low-cost point-of-care platforms, our data support the programmatic roll-out of CRP for pre-ART TB screening^3^, while better biomarkers are sought.

Our study has numerous strengths, including the size of the cohort, with a representative sample of >700 participants newly referred to initiate ART. The cohort were well-characterised and intensively investigated for TB, with 90% having two sputum culture results available, thus enabling a robust culture-based primary reference standard, complemented by sputum induction when required. In addition, sputum Ultra and urine diagnostics were systematically applied, and data linkage was performed to routinely collected data warehouse records to identify participants who were diagnosed and treated for TB following study enrolment. This enabled us to conduct secondary analyses using alternative reference standards, which supported the robustness of our primary findings. We implemented a laboratory and analysis pipeline using the Nanostring platform to measure seven candidate RNA signatures that have performed well in previous analyses^6–8^. The signatures were curated through our previous systematic review^6^ and were reproduced according to the original authors’ descriptions. The Nanostring pipeline demonstrated high levels of reproducibility and our head-to-head analysis showed superior discrimination for TB status when compared to RNA sequencing data in a subset of samples from our previously published presumptive TB cohort, thus reinforcing its robustness. Finally, CRP was also measured, enabling comparative head-to-head analyses with the candidate RNA biomarkers.

Our study is limited to a single centre, precluding assessments of generalisability across settings. This setting may be considered to be generally representative of populations with hyperendemic transmission of TB and HIV, but the relatively low specificity of the biomarkers means that the positive predictive value of these tests will reduce among PLHIV with lower prior probability of TB. Second, our targeted approach to RNA quantification precludes development of novel signatures beyond the 23 measured transcripts. Further genome-wide discovery will be required in such cohorts to identify novel biomarkers. Third, viral load was not available. We were therefore unable to test the hypothesis that high viral loads are independently associated with higher RNA signature scores. This limitation was mitigated by the availability of multiple other indices of HIV severity, which were associated with RNA signature scores, but failed to improve the performance of biomarkers in multivariable models.

In conclusion, RNA biomarkers showed better clinical utility to guide confirmatory TB testing for pre-ART screening than W4SS, but their performance did not exceed that of CRP, and fell short of WHO mandated targets. Interferon-independent approaches for host-response TB diagnostic screening may be required to improve specificity among PLHIV prior to ART initiation. Until then, the clinical and health-economic impact of widely available point-of-care CRP tests should be further evaluated for pre-ART TB screening.

## Data Availability

Processed Nanostring data will be provided as a supplementary file, along with accompanying metadata at the time of peer-reviewed publication.

## Footnotes

## Acknowledgements

The authors thank the Department of Health, Western Cape Data Warehouse team for providing TB testing and treatment data for patients for the period after study visit.

## Author contributions

Conceived and designed the study: RKG, GT, MN

Sample and clinical data collection: BR, GN, HM, BD, HT, ZP, SN, DM

Laboratory analysis: TM, AC, PK

Data analysis: TM, BR, RKG, CJC, AC, PK, GT, MN

Manuscript preparation: TM, RKG, BR, GT, MN with input from all authors

## References

1 World Health Organization. Global Tuberculosis Report 2022. 2022.

2 Gupta RK, Lucas SB, Fielding KL, Lawn SD. Prevalence of tuberculosis in post-mortem studies of HIV-infected adults and children in resource-limited settings. Aids 2015; 29: 1987–2002.

3 World Health Organization (WHO). Operational handbook on tuberculosis: Module 2: Systematic screening for tuberculosis disease. 2022 https://apps.who.int/iris/bitstream/handle/10665/340256/9789240022614-eng.pdf.

4 Dhana A, Hamada Y, Kengne AP, et al. Tuberculosis screening among ambulatory people living with HIV: a systematic review and individual participant data meta-analysis. Lancet Infect Dis 2022; 22: 507–18.

5 WHO. High-priority target product profiles for new tuberculosis diagnostics: report of a consensus meeting. 2014. https://www.who.int/tb/publications/tpp_report/en/ x(accessed May 21, 2019).

6 Gupta RK, Turner CT, Venturini C, et al. Concise whole blood transcriptional signatures for incipient tuberculosis: a systematic review and patient-level pooled meta-analysis. Lancet Respir Med 2020; 0. DOI:10.1016/S2213-2600(19)30282-6.

7 Turner CT, Gupta RK, Tsaliki E, et al. Blood transcriptional biomarkers for active pulmonary tuberculosis in a high-burden setting: a prospective, observational, diagnostic accuracy study. Lancet Respir Med 2020; 8: 407–19.

8 Mendelsohn SC, Mbandi SK, Fiore-gartland A, et al. Prospective multicentre head-to-head validation of host blood transcriptomic biomarkers for pulmonary tuberculosis by real-time PCR. Commun Med 2022. DOI:10.1038/s43856-022-00086-8.

9 Hoang LT, Jain P, Pillay TD, et al. Transcriptomic signatures for diagnosing tuberculosis in clinical practice: a prospective, multicentre cohort study. Lancet Infect Dis 2021; 21: 366–75.

10 Scriba TJ, Fiore-Gartland A, Penn-Nicholson A, et al. Biomarker-guided tuberculosis preventive therapy (CORTIS): a randomised controlled trial. Lancet Infect Dis 2021; 0. DOI:10.1016/s1473-3099(20)30914-2.

11 Sutherland JS, Spuy G Van Der, Gindeh A, et al. Diagnostic Accuracy of the Cepheid 3-gene Host Response Fingerstick Blood Test in a Prospective, Multi-site Study : Interim Results. 2022; 74.

12 Mendelsohn SC, Fiore-Gartland A, Penn-Nicholson A, et al. Validation of a host blood transcriptomic biomarker for pulmonary tuberculosis in people living with HIV: a prospective diagnostic and prognostic accuracy study. Lancet Glob Heal 2021; 9: e841–53.

13 Lawn SD, Kranzer K, Edwards DJ, McNally M, Bekker L-G, Wood R. Tuberculosis during the first year of antiretroviral therapy in a South African cohort using an intensive pretreatment screening strategy. AIDS 2010; 24: 1323–8.

14 Esmail H, Lai RP, Lesosky M, et al. Complement pathway gene activation and rising circulating immune complexes characterize early disease in HIV-associated tuberculosis. Proc Natl Acad Sci 2018; 115. DOI:10.1073/pnas.1711853115.

15 Turner CT, Brown J, Shaw E, et al. Persistent T Cell Repertoire Perturbation and T Cell Activation in HIV After Long Term Treatment. Front Immunol 2021; 12. DOI:10.3389/FIMMU.2021.634489.

16 Mulenga H, Musvosvi M, Mendelsohn SC, et al. Longitudinal Dynamics of a Blood Transcriptomic Signature of Tuberculosis. Am J Respir Crit Care Med 2021; 204: 1463–72.

17 Noursadeghi M, Gupta RK. New Insights into the Limitations of Host Transcriptional Biomarkers of Tuberculosis. Am J Respir Crit Care Med 2021; 204: 1363–5.

18 Cohen JF, Korevaar DA, Altman DG, et al. STARD 2015 guidelines for reporting diagnostic accuracy studies : explanation and elaboration. 2016; : 1–17.

19 Mutemaringa T, Heekes A, Boulle A, Tiffin N. Record linkage for Routinely Collected Health Data in an African Health Information Exchange. Int J Popul Data Sci 2022; 7. DOI:10.23889/IJPDS.V7I3.2022.

20 Krzanowski WJ, Hand DJ. ROC Curves for Continuous Data. Chapman and Hall/CRC, 2009 DOI:10.1201/9781439800225.

21 Gupta RK, Turner CT, Venturini C, et al. Concise whole blood transcriptional signatures for incipient tuberculosis: a systematic review and patient-level pooled meta-analysis. Lancet Respir Med 2020; 0. DOI:10.1016/S2213-2600(19)30282-6.

22 Penn-Nicholson A, Mbandi SK, Thompson E, et al. RISK6, a 6-gene transcriptomic signature of TB disease risk, diagnosis and treatment response. Sci Rep 2020; 10: 1–21.

23 Pollara G, Turner CT, Rosenheim J, et al. Exaggerated IL-17A activity in human in vivo recall responses discriminates active tuberculosis from latent infection and cured disease. Sci Transl Med 2021; 13. DOI:10.1126/scitranslmed.abg7673.

24 Leek J, Johnson W, Parker H, et al. sva: Surrogate Variable Analysis. 2019.

25 DeLong ER, DeLong DM, Clarke-Pearson DL. Comparing the areas under two or more correlated receiver operating characteristic curves: a nonparametric approach. Biometrics 1988; 44: 837–45.

26 Brown M. rmda: Risk Model Decision Analysis. 2018.

27 Calderwood CJ, Reeve BW, Mann T, et al. Clinical utility of C-reactive protein-based triage for presumptive pulmonary tuberculosis in South African adults. J Infect 2022; published online Nov. DOI:10.1016/j.jinf.2022.10.041.

28 Kaforou M, Wright VJ, Oni T, et al. Detection of tuberculosis in HIV-infected and -uninfected African adults using whole blood RNA expression signatures: a case-control study. PLoS Med 2013; 10: e1001538.

29 Rudolf F. The Bandim TBscore – reliability, further development, and evaluation of potential uses. Glob Health Action 2014; 7: 24303.

