## Supplementary Tables and Figures for "Blood RNA biomarkers for tuberculosis screening in people living with HIV prior to anti-retroviral therapy initiation: A diagnostic accuracy study"

**Supplementary Table 1**

***Approach to calculation of blood RNA biomarker scores***

All calculations are performed on log-2 transformed data. A single Nanostring probe was used to measure FCGR1A and FCGR1B.

Commented [GR1]: @Maddy - check wording
Commented [NM2R1]: fine

| Signature | Calculation |
| --- | --- |
| *BATF2*[1] | BATF2 |
| *Gliddon3*[2] | (FCGR1A + C1QB) - (ZNF296) |
| *RISK6*[3] | ((GBP2 + FCGR1B + SERPING1)/3) - ((TUBGCP6 + TRMT2A + SDR39U1)/3)) |
| *Roe3*[4] | (BATF2 + SCARF1 + GBP5)/3 |
| *Suliman4*[5] | (GAS6 + SEPT4) - (CD1C + BLK) |
| *Sweeney3*[6] | (GBP5 + DUSP3)/2) - KLF2 |
| *Zak11*[7] | Support vector machine model trained on original training dataset[8] |

**Supplementary Table 2**

***Baseline characteristics of the study cohort, stratified by inclusion***

| Characteristic | Overall, N = 862 | Included, N = 707[1] | Excluded, N = 155[1] | p-value[2] |
|---|---|---|---|---|
| Age (years) | 32 (26, 39) | 32 (27, 39) | 32 (25, 38) | 0.2 |
| Gender | | | | 0.6 |
| Female | 501 (58%) | 407 (58%) | 94 (61%) | |
| Male | 360 (42%) | 299 (42%) | 61 (39%) | |
| Missing | 1 | 1 | 0 | |
| Previous TB | 124 (14%) | 98 (14%) | 26 (17%) | 0.4 |
| CD4 (cells/mm3) | 299 (172, 486) | 306 (184, 486) | 274 (129, 490) | 0.12 |
| Missing | 8 | 5 | 3 | |
| CD4 <200 cells/mm3 | 248 (29%) | 193 (27%) | 55 (36%) | 0.041 |
| Missing | 8 | 5 | 3 | |
| Haemoglobin (g/dl) | 12.70 (11.30, 13.90) | 12.70 (11.30, 13.90) | 12.50 (11.25, 13.83) | 0.4 |
| Missing | 177 | 150 | 27 | |
| Body mass index (kg/m2) | 24 (21, 29) | 24 (21, 29) | 23 (20, 29) | 0.4 |
| Missing | 2 | 2 | 0 | |
| Middle upper arm circumference (cm) | 27.0 (25.0, 30.0) | 27.0 (25.0, 30.0) | 27.0 (25.0, 30.5) | 0.7 |
| WHO 4-symptom screen positive | 487 (56%) | 406 (57%) | 81 (52%) | 0.3 |
| TBscoreII | 1.00 (0.00, 2.00) | 1.00 (0.00, 2.00) | 1.00 (0.00, 1.25) | 0.6 |
| Missing | 39 | 36 | 3 | |
| CRP (mg/L) | 6 (2, 32) | 6 (2, 32) | 6 (2, 34) | 0.6 |
| Missing | 17 | 0 | 17 | |
| Number of valid sputum cultures | | | | 0.7 |
| 0 | 37 (4.3%) | 31 (4.4%) | 6 (3.9%) | |
| 1 | 50 (5.8%) | 43 (6.1%) | 7 (4.5%) | |
| 2 | 775 (90%) | 633 (90%) | 142 (92%) | |
| Sputum culture positive | 107 (13%) | 89 (13%) | 18 (12%) | 0.8 |
| Missing | 37 | 31 | 6 | |
| Sputum Ultra | | | | 0.4 |
| Negative | 754 (89%) | 616 (88%) | 138 (91%) | |
| Trace | 19 (2.2%) | 18 (2.6%) | 1 (0.7%) | |
| Positive | 78 (9.2%) | 65 (9.3%) | 13 (8.6%) | |
| Missing | 11 | 8 | 3 | |
| Urine LAM | 24 (2.8%) | 18 (2.6%) | 6 (3.9%) | 0.4 |
| Missing | 5 | 3 | 2 | |
| Urine Ultra | 45 (5.3%) | 37 (5.3%) | 8 (5.3%) | >0.9 |
| Missing | 10 | 6 | 4 | |
| Sputum culture or Ultra positive | 113 (13%) | 94 (13%) | 19 (12%) | 0.9 |
| Missing | 12 | 9 | 3 | |
| Any positive TB test | 137 (16%) | 112 (16%) | 25 (16%) | >0.9 |
| Missing | 11 | 8 | 3 | |

| Characteristic | Overall, N = 862 | Included, N = 707[1] | Excluded, N = 155[1] | p-value[2] |
|---|---|---|---|---|
| Recorded TB diagnosis or treatment within 6 months | 152 (18%) | 130 (18%) | 22 (14%) | 0.3 |

[1]Statistics presented: median (IQR); n (%)

[2]Statistical tests performed: Wilcoxon rank-sum test; chi-square test of independence; Fisher's exact test

**Supplementary Table 3**

***Linear regression model showing factors associated with higher Suliman4 scores***

| Characteristic | Beta | 95% CI[1] | p-value |
|---|---|---|---|
| (Intercept) | 3.8 | 2.2, 5.4 | <0.001 |
| Age (years) | 0.01 | 0.00, 0.03 | 0.2 |
| Gender | | | |
| Female | — | — | |
| Male | 0.13 | -0.23, 0.49 | 0.5 |
| CD4 count (cells/mm3; per 10 unit increase) | -0.02 | -0.03, -0.02 | <0.001 |
| Haemoglobin (g/dl) | -0.24 | -0.32, -0.16 | <0.001 |
| Body mass index (kg/m2) | -0.01 | -0.04, 0.01 | 0.4 |
| Respiratory rate (per min) | 0.07 | 0.02, 0.11 | 0.004 |
| Number of W4SS symptoms | 0.40 | 0.26, 0.55 | <0.001 |
| Sputum culture positive | 1.5 | 1.0, 2.0 | <0.001 |

[1]CI = Confidence Interval

**Supplementary Table 4**

***Table showing thresholds where screening approaches have higher net benefit than confirmatory testing for all and none***

Thresholds shown as threshold probabilities and as number willing to test ranges with confirmatory tests per true TB case detected.

| Score | Threshold probability range | NWT range |
|---|---|---|
| CRP >= 5mg/L & Suliman4 >= Z2 | 0.04 - 0.29 | 3.5 - 23.8 |
| Suliman4 >= Z2 | 0.04 - 0.23 | 4.3 - 23.8 |
| CRP >= 5mg/L | 0.04 - 0.2 | 5 - 22.2 |
| W4SS | 0.06 - 0.18 | 5.5 - 15.4 |

### Supplementary Table 5

***Table showing greedy forward search looking for an optimised signature***

The full dataset was temporally split into 75%/25% development and validation sets. Increasing numbers of genes were iteratively added, in order of their discrimination for TB as single predictors.

| Number of genes | Added gene | Logistic regression | Support vector machine | Disease risk score | Difference in geometric means |
|---|---|---|---|---|---|
| 1 | SEPT4 | 0.77 (0.66 - 0.88) | 0.77 (0.66 - 0.88) | 0.77 (0.66 - 0.88) | 0.77 (0.66 - 0.88) |
| 2 | FCGR1A | 0.77 (0.66 - 0.87) | 0.73 (0.63 - 0.83) | 0.77 (0.66 - 0.87) | 0.77 (0.66 - 0.87) |
| 3 | SERPING1 | 0.76 (0.66 - 0.87) | 0.75 (0.65 - 0.85) | 0.76 (0.65 - 0.86) | 0.76 (0.65 - 0.86) |
| 4 | BATF2 | 0.76 (0.65 - 0.86) | 0.75 (0.64 - 0.85) | 0.75 (0.65 - 0.86) | 0.75 (0.65 - 0.86) |
| 5 | CD1C | 0.75 (0.63 - 0.87) | 0.5 (0.4 - 0.61) | 0.76 (0.65 - 0.87) | 0.75 (0.64 - 0.86) |
| 6 | DUSP3 | 0.76 (0.64 - 0.87) | 0.71 (0.59 - 0.83) | 0.76 (0.65 - 0.87) | 0.75 (0.63 - 0.86) |
| 7 | C1QB | 0.75 (0.63 - 0.86) | 0.73 (0.62 - 0.84) | 0.76 (0.65 - 0.86) | 0.75 (0.63 - 0.86) |
| 8 | KLF2 | 0.76 (0.65 - 0.87) | 0.71 (0.6 - 0.82) | 0.76 (0.65 - 0.87) | 0.76 (0.65 - 0.87) |
| 9 | ZNF296 | 0.78 (0.67 - 0.88) | 0.77 (0.67 - 0.87) | 0.76 (0.65 - 0.87) | 0.77 (0.66 - 0.88) |
| 10 | GBP1 | 0.78 (0.68 - 0.88) | 0.75 (0.64 - 0.85) | 0.76 (0.65 - 0.87) | 0.77 (0.66 - 0.88) |
| 11 | GAS6 | 0.77 (0.67 - 0.88) | 0.74 (0.63 - 0.86) | 0.77 (0.66 - 0.88) | 0.77 (0.66 - 0.88) |
| 12 | GBP5 | 0.8 (0.71 - 0.89) | 0.76 (0.66 - 0.87) | 0.77 (0.66 - 0.87) | 0.77 (0.66 - 0.88) |
| 13 | SDR39U1 | 0.8 (0.71 - 0.89) | 0.76 (0.66 - 0.86) | 0.77 (0.66 - 0.88) | 0.77 (0.66 - 0.89) |
| 14 | SCARF1 | 0.78 (0.69 - 0.88) | 0.74 (0.64 - 0.85) | 0.76 (0.65 - 0.87) | 0.77 (0.66 - 0.88) |
| 15 | TRMT2A | 0.79 (0.69 - 0.88) | 0.74 (0.64 - 0.85) | 0.77 (0.66 - 0.88) | 0.77 (0.66 - 0.88) |
| 16 | ETV7 | 0.78 (0.68 - 0.88) | 0.74 (0.63 - 0.85) | 0.76 (0.65 - 0.87) | 0.77 (0.66 - 0.88) |
| 17 | TAP1 | 0.78 (0.68 - 0.88) | 0.74 (0.64 - 0.85) | 0.76 (0.65 - 0.87) | 0.77 (0.66 - 0.88) |
| 18 | GBP2 | 0.78 (0.68 - 0.88) | 0.74 (0.63 - 0.84) | 0.76 (0.65 - 0.87) | 0.77 (0.66 - 0.88) |
| 19 | BLK | 0.78 (0.68 - 0.87) | 0.74 (0.64 - 0.85) | 0.76 (0.65 - 0.87) | 0.76 (0.65 - 0.87) |
| 20 | TUBGCP6 | 0.78 (0.68 - 0.87) | 0.75 (0.64 - 0.85) | 0.76 (0.65 - 0.87) | 0.77 (0.66 - 0.88) |
| 21 | STAT1 | 0.77 (0.68 - 0.87) | 0.74 (0.63 - 0.85) | 0.76 (0.65 - 0.87) | 0.77 (0.66 - 0.88) |
| 22 | TRAFD1 | 0.78 (0.69 - 0.88) | 0.76 (0.66 - 0.86) | 0.76 (0.65 - 0.87) | 0.76 (0.65 - 0.87) |

**Supplementary Figure 1**

***Sample size calculations***

Relationship between total sample size required and proportion of positive cases within the sample stratified by target area under the receiver operating characteristic curve (AUROC) to discriminate between cases and controls with 95% lower bound confidence interval of 0.05 from target AUROC. Dashed line represents sample size of present study (N=707).

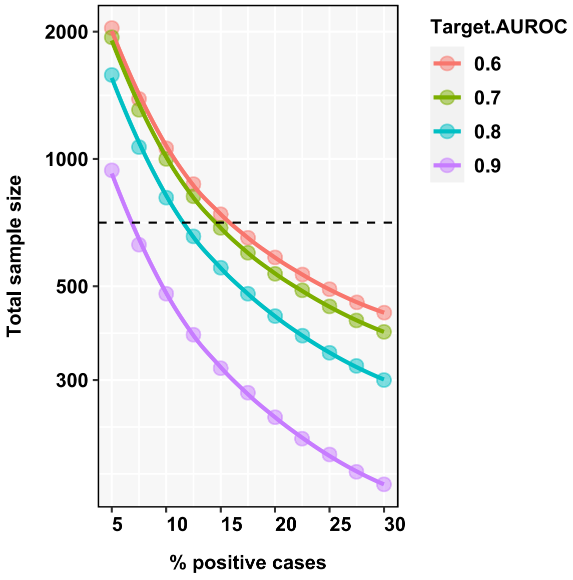

**Supplementary Figure 2**

***Coefficients of variation for reference RNA samples, stratified by Nanostring codeset batch.***

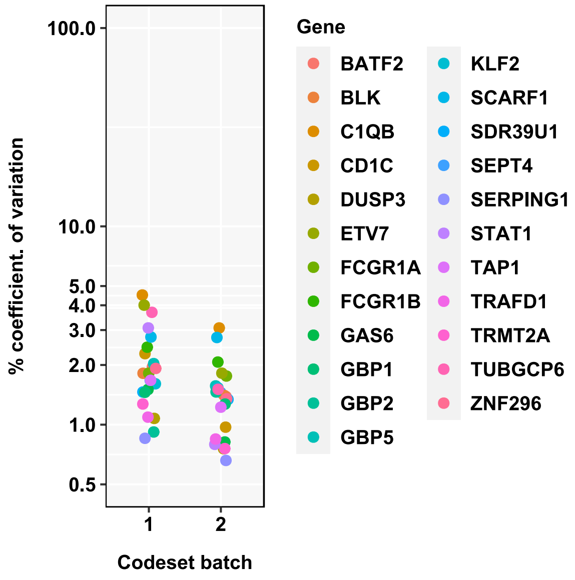

**Supplementary Figure 3**

***Discrimination of TB signatures in BAR-TB dataset using Nanostring versus RNAseq (n=59).***

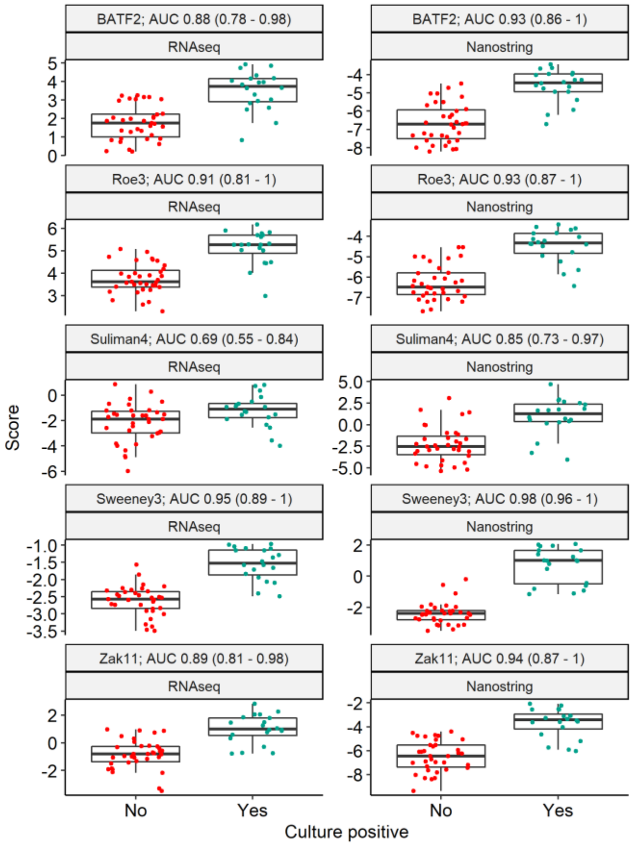

**Supplementary Figure 4**

***PCA of reference RNA samples***

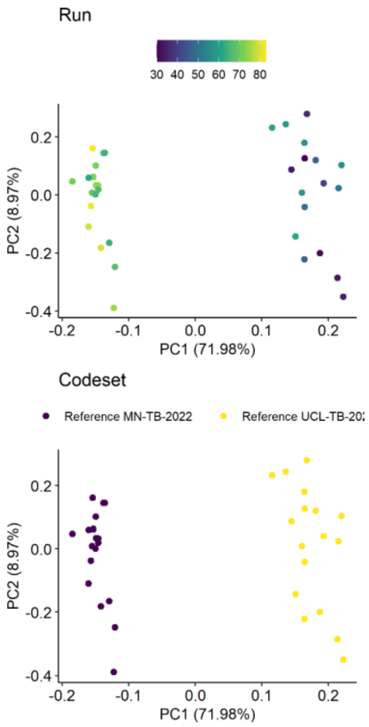

**Supplementary Figure 5**

***Distribution of gene expression values normalized to GAPDH by Nanostring codeset batch***

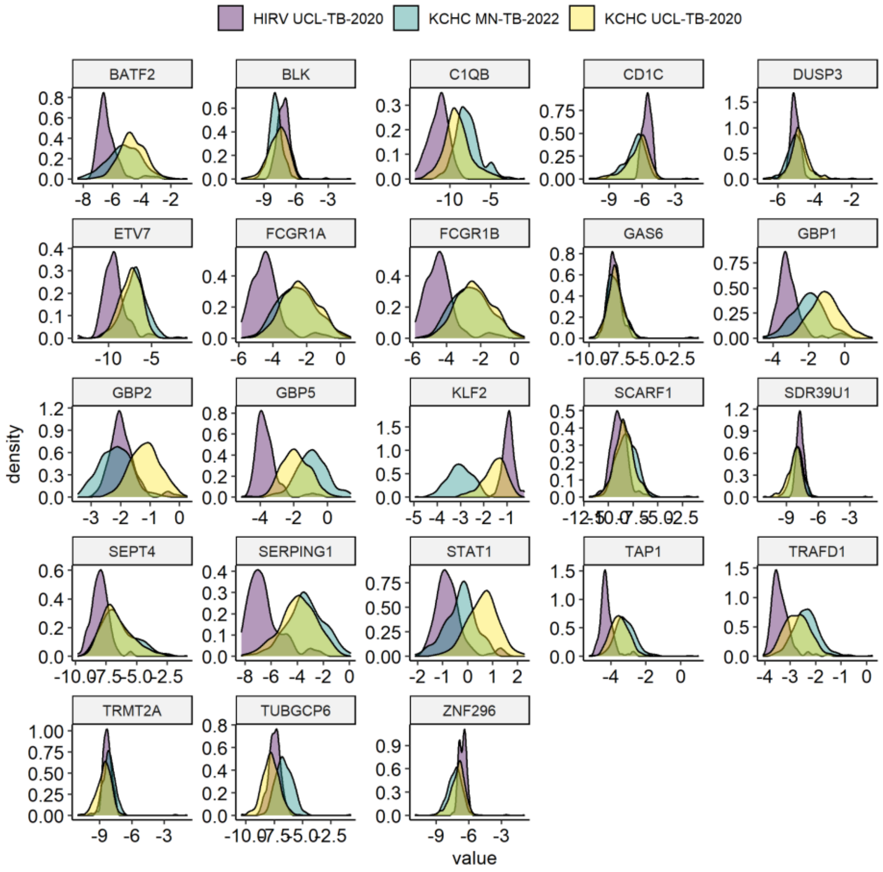

**Supplementary Figure 6**

***Distribution of gene expression values normalized to GAPDH by Nanostring codeset batch after COMBAT correction***

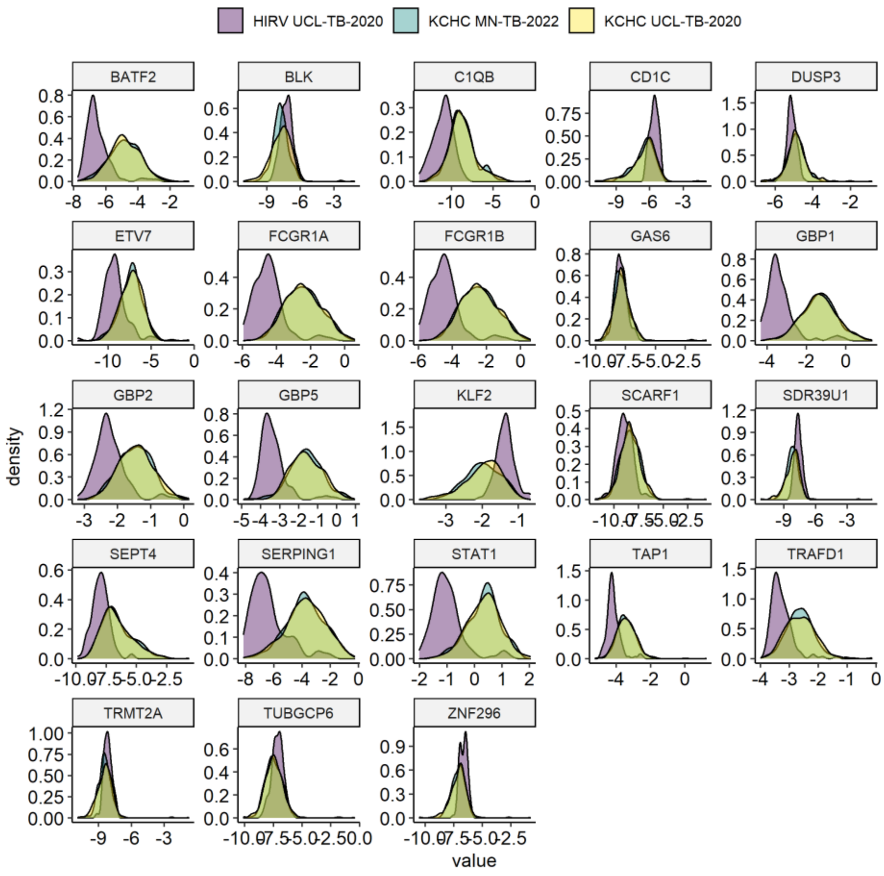

**Supplementary Figure 7**

***Distribution of gene expression values normalized to GAPDH by Nanostring codeset batch after reference RNA normalization***

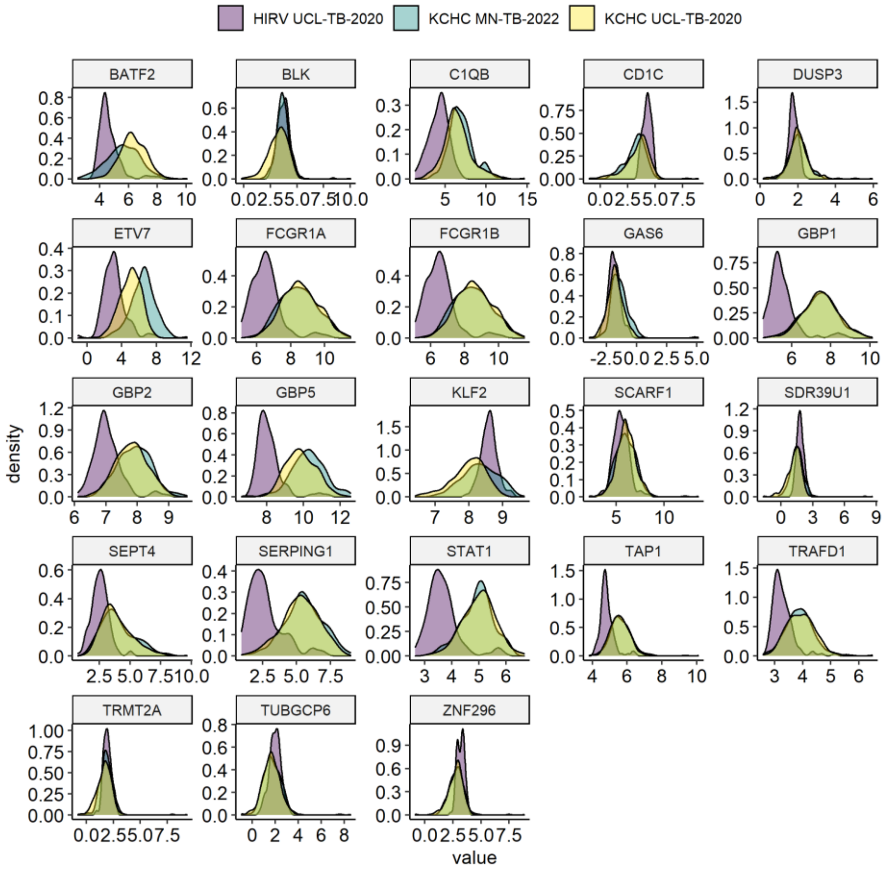

**Supplementary Figure 8**

***Discrimination of Roe3 and Zak11 signatures for (A) incipient and (B) presumptive TB using simple calculations (geometric means), compared to scaled and unscaled SVMs***

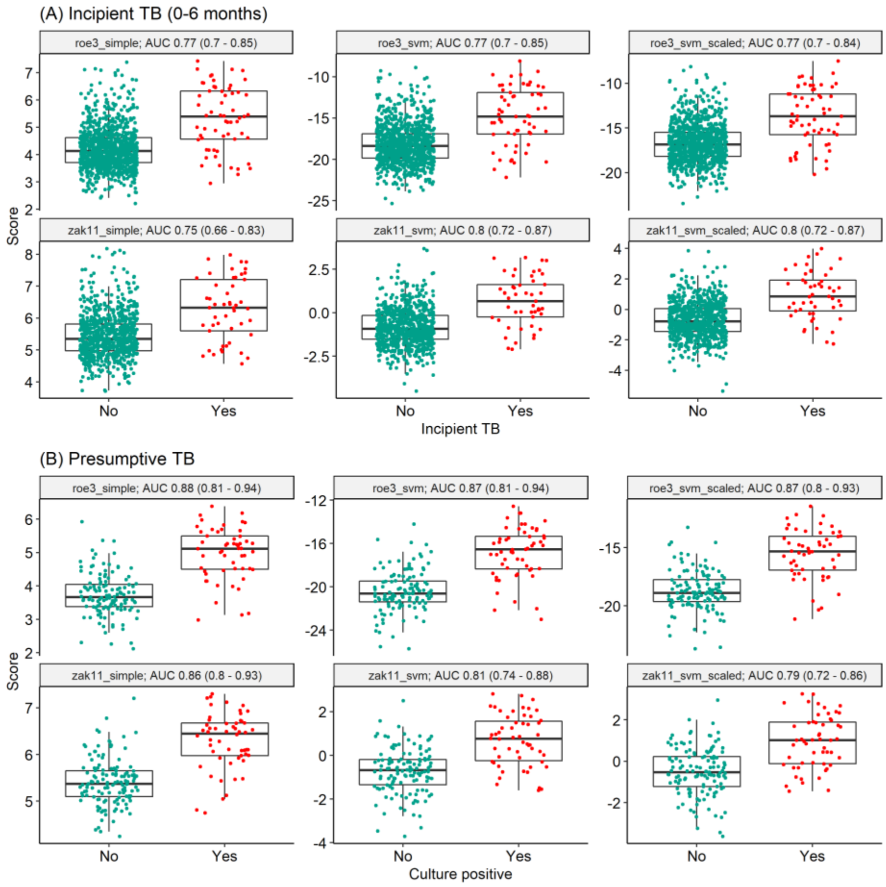

**Supplementary Figure 9**

***Interval time between enrolment to the study and first TB diagnosis or treatment initiation recorded in study or registry data.***

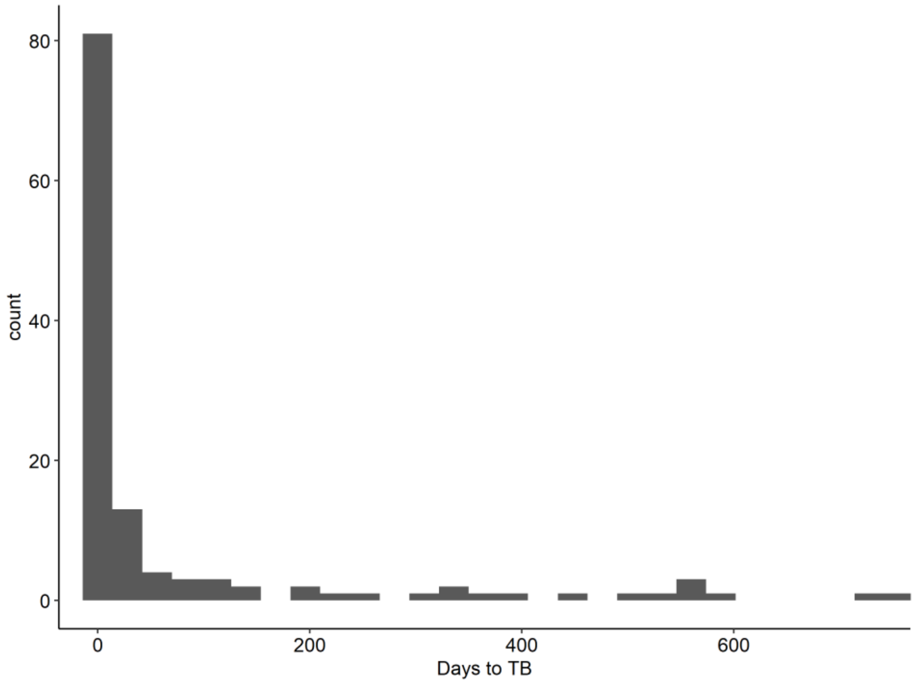

**Supplementary Figure 10**

***Co-correlation of blood RNA biomarkers and CRP***

Spearman rank co-correlation matrix of blood RNA biomarkers and CRP (n=707), with hierarchical clustering using complete linkage method.

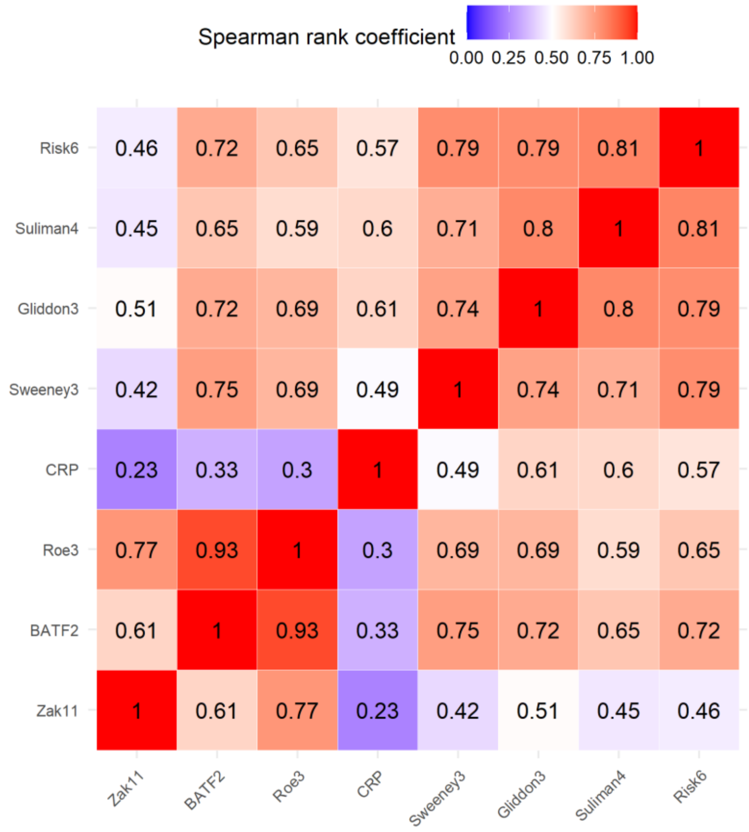

**Supplementary Figure 11**

***Associations between Suliman4 blood RNA biomarker Z-score sand indices of HIV/TB disease severity***

Individual scatter plots of Suliman4 blood RNA biomarker Z score with selected variables (indicated) associated with HIV and/or TB disease burden/severity, for all participant data (N=676) stratified by sputum Mtb culture result. Spearman correlation coefficients and p values are shown for each pairwise analysis. Sputum smear grade is shown as 1 (scanty), 2 (+), 3 (++) and 4 (+++)..

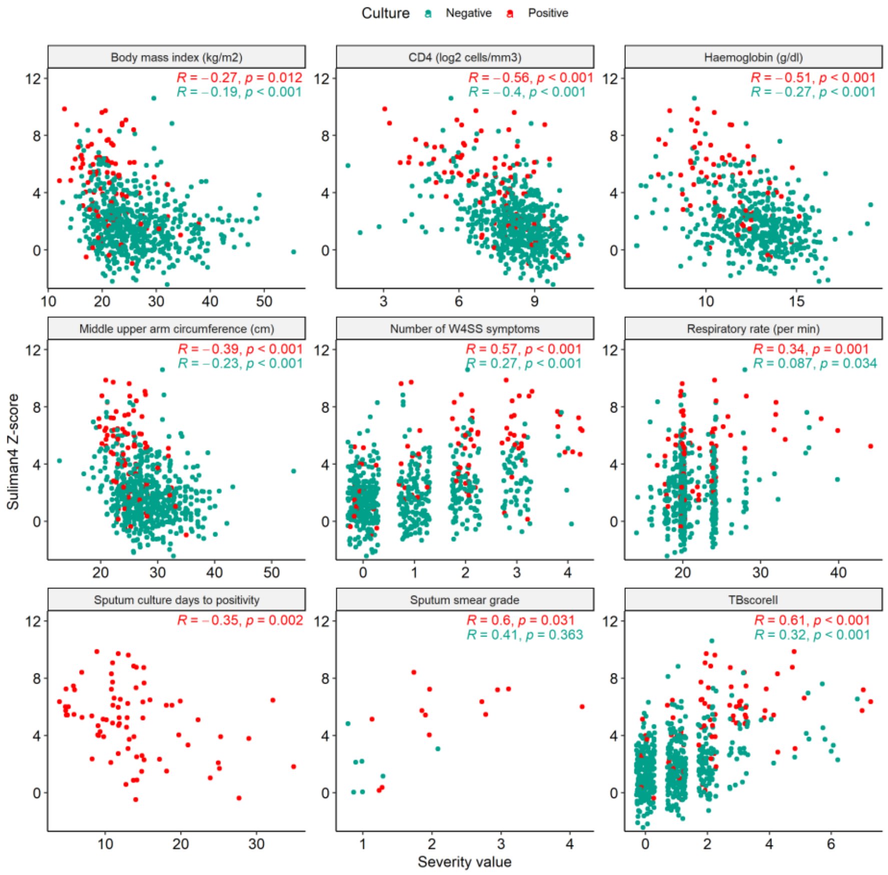

**Supplementary Figure 12**

***Linear regression model of predictors for Suliman4 scores***

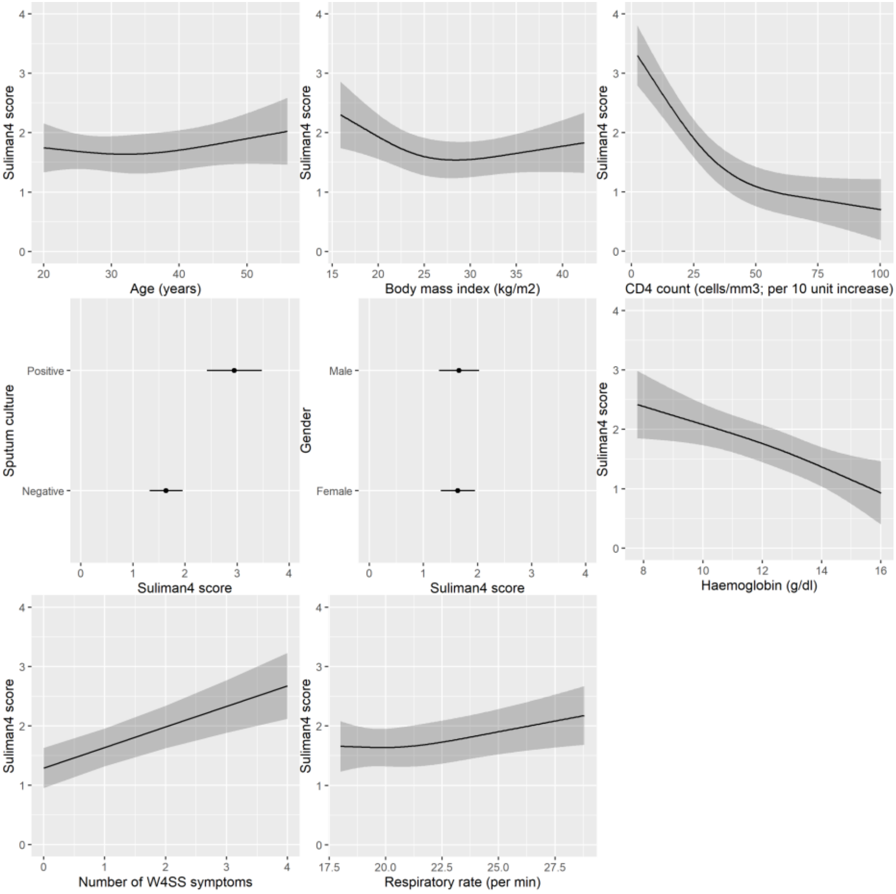

**Supplementary Figure 13**

***Greedy forward search looking for an optimised signature***

The full dataset was temporally split into 75%/25% development and validation sets. Increasing numbers of genes were iteratively added, in order of their discrimination for TB as single predictors.

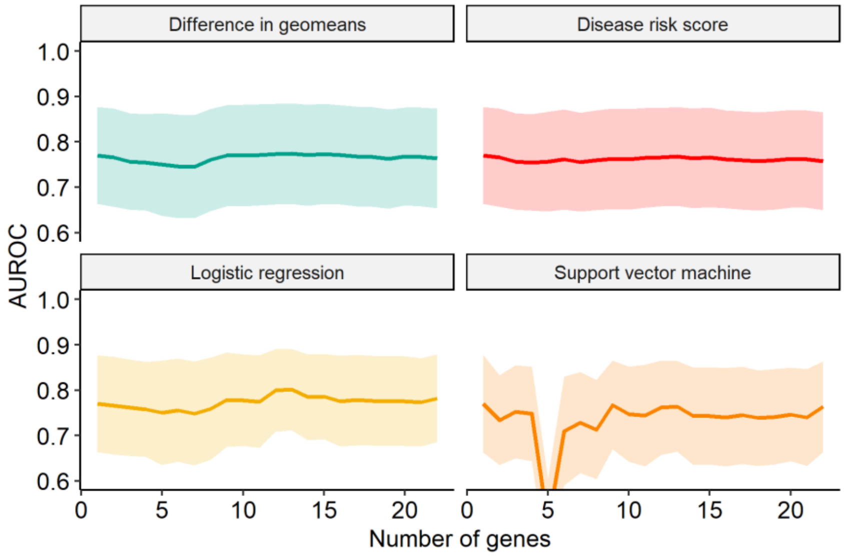

**Supplementary Figure 14**

***Sensitivity analysis: Sputum culture or Ultra positivity***

Scores and discrimination of RNA signatures for secondary outcome of sputum culture or Ultra positivity (n = 698 participants). Scores are shown as Z-scores for RNA signatures, and log-2 transformed CRP (mg/L). Discrimination presented as area under the receiver operating characteristic curve (AUC), with 95% confidence intervals.

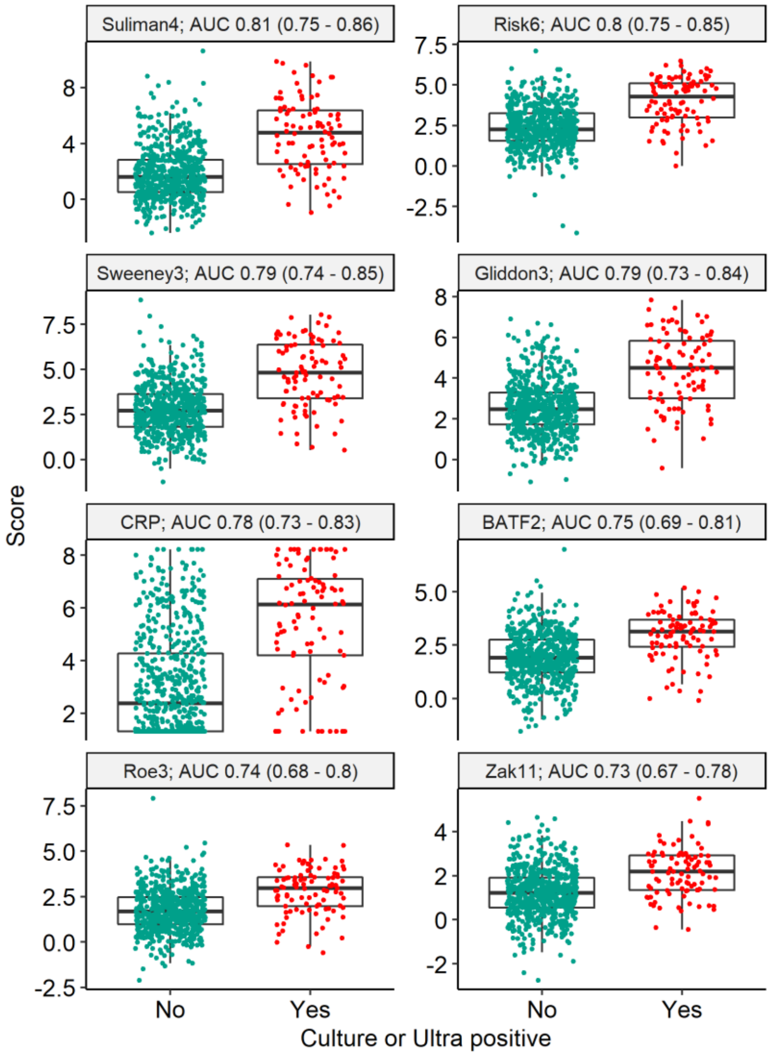

**Supplementary Figure 15**

***Sensitivity analysis: Any positive TB test***

Scores and discrimination of RNA signatures for secondary outcome of any positive TB test, including urine LAM and urine Ultra (n = 699 participants). Scores are shown as Z-scores for RNA signatures, and log-2 transformed CRP (mg/L). Discrimination presented as area under the receiver operating characteristic curve (AUC), with 95% confidence intervals.

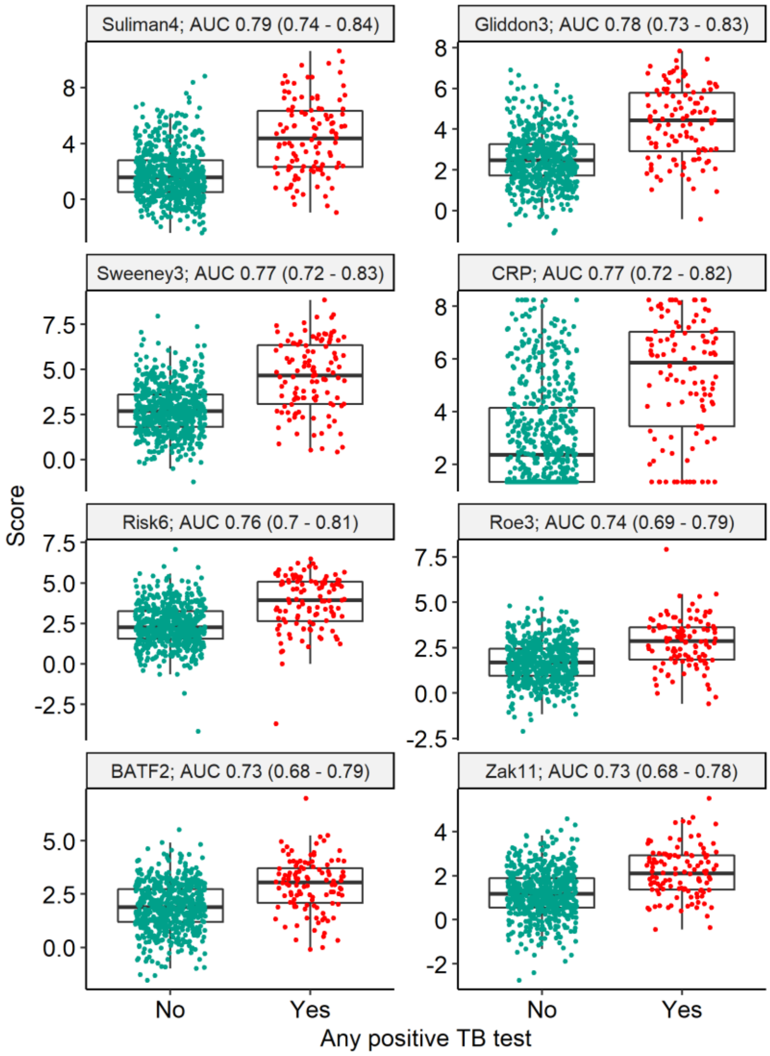

**Supplementary Figure 16**

***Sensitivity analysis: Recorded TB diagnosis or treatment within 6 months***

Scores and discrimination of RNA signatures for secondary outcome of recorded TB diagnosis or treatment within 6 months (n = 707 participants). Scores are shown as Z-scores for RNA signatures, and log-2 transformed CRP (mg/L). Discrimination presented as area under the receiver operating characteristic curve (AUC), with 95% confidence intervals.

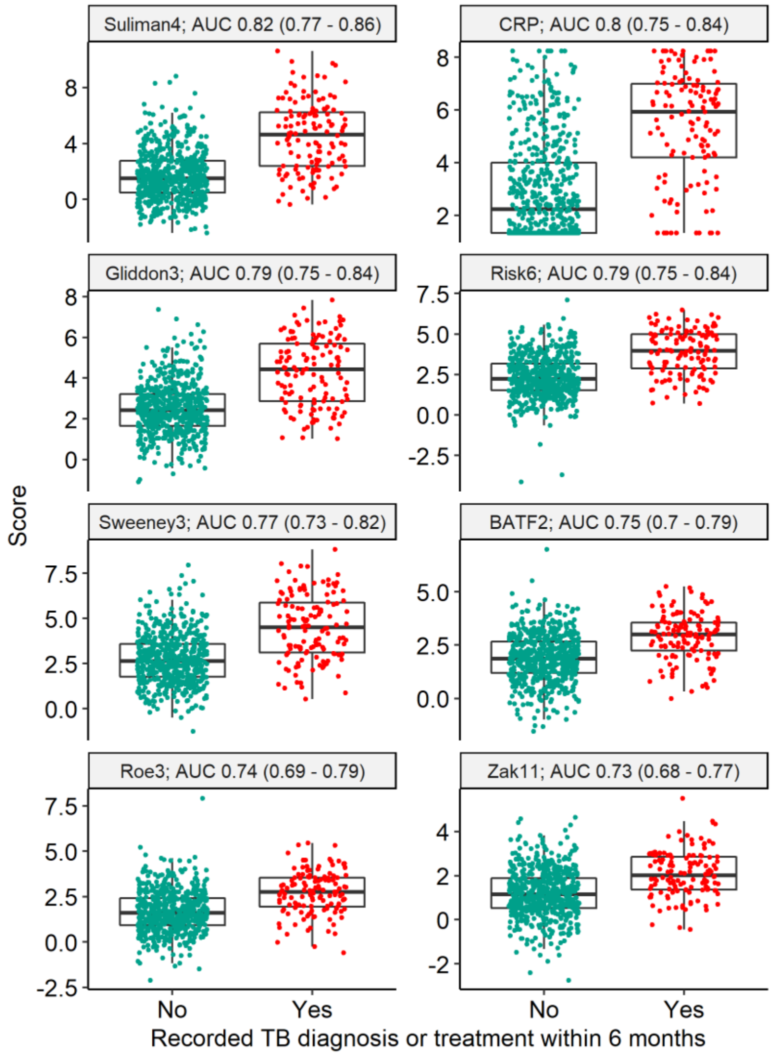

**Supplementary Figure 17**

***Sensitivity analysis: Using reference RNA-normalised data***

Scores and discrimination of RNA signatures for primary outcome of sputum culture (n = 676 participants), using reference RNA normalisation of Nanostring data as sensitivity analysis. Scores are shown as untransformed scores for RNA signatures, and log-2 transformed CRP (mg/L). Discrimination presented as area under the receiver operating characteristic curve (AUC), with 95% confidence intervals.

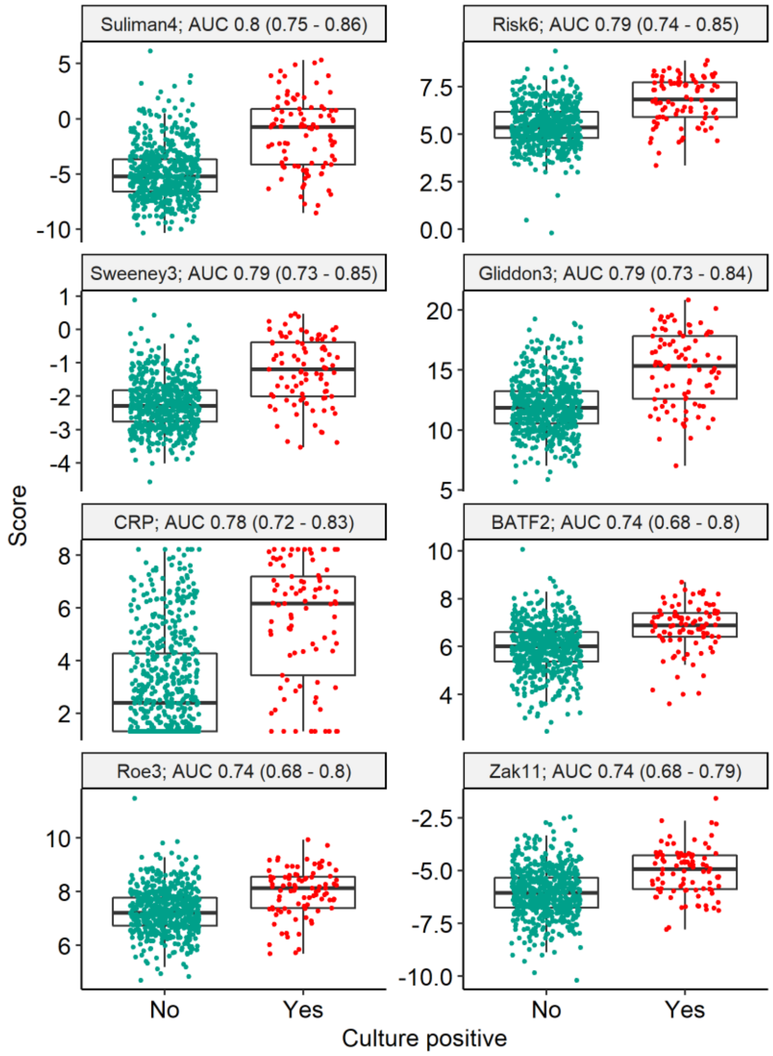

### References

1 Roe JK, Thomas N, Gil E, *et al.* Blood transcriptomic diagnosis of pulmonary and extrapulmonary tuberculosis. *JCI insight* 2016; **1**: e87238.

2 Gliddon HD, Kaforou M, Alikian M, *et al.* Identification of reduced host transcriptomic signatures for tuberculosis and digital PCR-based validation and quantification. *bioRxiv* 2019; : 583674.

3 Penn-Nicholson A, Mbandi SK, Thompson E, *et al.* RISK6, a 6-gene transcriptomic signature of TB disease risk, diagnosis and treatment response. *Sci Rep* 2020; **10**: 1–21.

4 Roe J, Venturini C, Gupta RK, *et al.* Blood transcriptomic stratification of short-term risk in contacts of tuberculosis. *Clin Infect Dis* 2019; published online March 28. DOI:10.1093/cid/ciz252.

5 Suliman S, Thompson E, Sutherland J, *et al.* Four-gene Pan-African Blood Signature Predicts Progression to Tuberculosis. *Am J Respir Crit Care Med* 2018; **197**: 1198–208.

6 Sweeney TE, Braviak L, Tato CM, Khatri P. Genome-wide expression for diagnosis of pulmonary tuberculosis: a multicohort analysis. *Lancet Respir Med* 2016; **4**: 213–24.

7 Darboe F, Mbandi SK, Thompson EG, *et al.* Diagnostic performance of an optimized transcriptomic signature of risk of tuberculosis in cryopreserved peripheral blood mononuclear cells. *Tuberculosis* 2018; **108**: 124–6.

8 Zak DE, Penn-Nicholson A, Scriba TJ, *et al.* A blood RNA signature for tuberculosis disease risk: a prospective cohort study. *Lancet* 2016; **387**: 2312–22.
